# Mixed ctDNA dynamics and decreased detection rates in early-stage lung cancer patients during radiation treatment

**DOI:** 10.1101/2024.06.03.24308298

**Authors:** Christopher Boniface, Kathryn Baker, Christopher Deig, Carol Halsey, Taylor Kelley, Ramtin Rahmani, Garth Tormoen, Paul T Spellman, Nima Nabavizadeh

## Abstract

Quantification and detection of circulating tumor DNA (ctDNA) has been used to identify the presence of cancers. Ablative radiation therapy kills tumor cells to reduce tumor burden and it follows that these dying tumor cells could lead to increased ctDNA abundance. We carried out deep, error-corrected sequencing of cell-free DNA collected serially from 12 stage I, and 2 stage II/III non-small cell lung cancer (NSCLC) patients undergoing external-beam radiation treatment (EBRT) after initial diagnosis. We found that ctDNA detection rates decreased at the first blood draw as compared to baseline (43% to 7% of patients). Total ctDNA abundance decreased in 6 patients and increased in 5 patients between those same blood draws, with one patient showing evidence of tumoral heterogeneity. Both patients with stage II/III disease had the largest increases in ctDNA abundance from baseline. Multiple blood draws improved ctDNA detection rates from 43% to 50% with a second blood draw and to 71% with 4 blood draws. Additionally, EGFR mutations were detectable in 6 patients during EBRT that were not detected prior to treatment. Taken together, these results provide an early-stage NSCLC counterpoint to previous work that reported improved ctDNA detection after radiation therapy in more advanced disease.

## 1. Introduction

Non-small cell lung cancer (NSCLC) constitutes 20% of new cancer cases and nearly one quarter of all cancer deaths in the US [1]. Early detection and intervention significantly improve patient outcome and long-term survival, yet over 75% of patients have regional or metastatic disease at the time of diagnosis. Recent efforts to expand access to low-dose CT screening for at-risk populations has been shown to improve survival rates by detecting cancer at earlier stages, but this comes with significant risk of increased over-diagnosis and over-treatment [2, 3].

Tumor mutation profiling in NSCLC is also becoming more valuable as new treatments are approved for genetic alterations in genes such as EGFR. These so-called ‘actionable’ mutations can confer resistance to first line treatment and become dominant in the primary tumor and metastases at later stages [4–6]. Although the clinical use of liquid biopsy is expanding, molecular-based treatment guidance has traditionally relied on solid-tissue biopsies for tumor profiling. Yet, solid-tissue biopsy is invasive, risky, and often insufficient or impossible in early-stage NSCLC patients. Consequently, many patients suspected of early-stage NSCLC are treated based on imaging diagnosis alone, without biopsy conformation of suspicious lung nodules. Liquid biopsy is therefore uniquely suited as a companion diagnostic to CT screening and in cases where biopsy confirmation of an imaging-based diagnosis is not available. It is also useful for tumor mutation profiling in leu of a solid-tissue biopsy. However, such an assay remains challenging to implement due to the low fractional abundance of tumor-derived biomarkers, such as circulating tumor DNA (ctDNA), in early-stage NSCLC.

ctDNA detection rates in stage I NSCLC patients range between 25-60% using next-generation sequencing (NGS) of commonly mutated genes [7–9]. Early-stage tumors (∼1 cm^3^) are estimated to contribute as little as 1-2 haploid genome equivalents per 15 ml plasma or <0.1% of total cell-free DNA, which is below the error-rate of traditional NGS [10, 11]. Molecular barcoding and computational strategies have been developed to discriminate between tumor-derived mutations and background-error in NGS [12, 13]. However, identifying tumor-derived ctDNA at mutant allele fractions (MAFs) below 0.1%-1% without prior genotyping of the tumor continues to be unreliable and presents unique challenges. Furthermore, as cell-free DNA is thought to be primarily derived from white blood cells (WBCs), somatic mutations derived from clonal hematopoiesis (CH) can confound ctDNA detection [14].

Our lab and others have observed increased ctDNA abundance after exposure to ionizing radiation in animal models and in cancer patients [15–20]. We hypothesized that ablative doses of radiation, such as those used in hypofractionated external-beam radiation treatment (EBRT), would lead to increased ctDNA shedding due to cytotoxicity and could therefore improve the detection rates of ctDNA-based assays in early-stage NSCLC. Furthermore, this increase in ctDNA shedding could improve liquid biopsy-based tumor genotyping. The routine radiation treatment of patients presenting suspicious lung nodules at our institution presented an opportunity to test this hypothesis. We sought to leverage the possible ctDNA enrichment effects of EBRT with computation-based error reduction workflows to improve ctDNA detection and characterization.

## 2. Materials and Methods

### 2.1 NSCLC patient and healthy control study consent, treatment, and sample acquisition

Human specimens and data (including whole blood and clinical information) were prospectively acquired from participants undergoing first-line external-beam radiation after diagnosis of NSCLC by imaging (n = 14, 12 x stage I, 1 x stage II, and 1 x stage III) after their informed written consent (Oregon Health & Science University Institutional Review Board Study #10163, first approved 19 October 2017). All patients were treated with 48-60 Gy external-beam radiation in 4-8 fractions (Table 1 and Fig. 1). Blood samples were collected in EDTA tubes prior to treatment (baseline, BL), within 21-192 (median = 47) hours following at least 1 treatment (Tx1), and at various time during EBRT. Three patients had draws following the completion of treatment at 48- to 96-hour intervals (patients tb184, tb187, and tb196). Blood was collected from 10 healthy individuals between 25-55 years of age after their informed written consent under the same IRB.

**Fig. 1.**
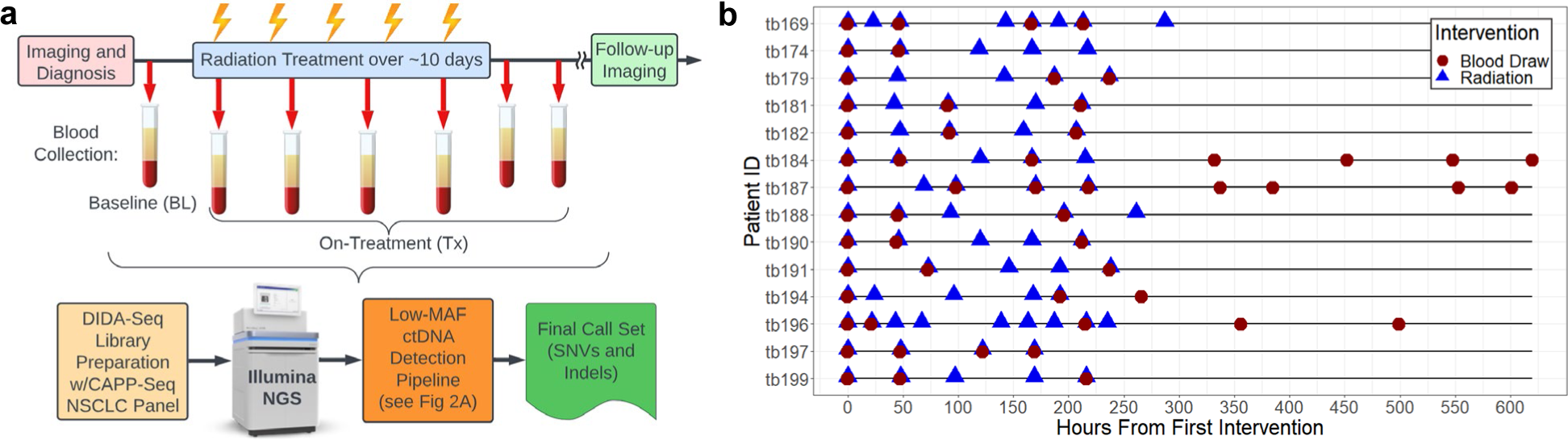
Idealized sample collection, and data analysis schematic and actual cohort treatment and blood draw timelines. An idealized timeline for patient blood sample collection during external beam radiation treatment (EBRT) is shown (**a**) as well as the general workflow for library preparation, sequencing, and analysis (**b**). EBRT and blood collection timing and number varied between patients. Post-treatment blood collection occurred in 3 patients and follow-up imaging varied from 6 to 8 weeks following the final EBRT fraction. [DIDA-Seq, dual-indexed degenerate adaptor sequencing [36]; CAPP-Seq, cancer personalized profiling by deep sequencing [9]); NSCLC, non-small cell lung cancer; NGS, next-generation sequencing; MAF, mutant allele fraction; SNV, single nucleotide variant; Indel, insertion or deletion].

**Table 1.**
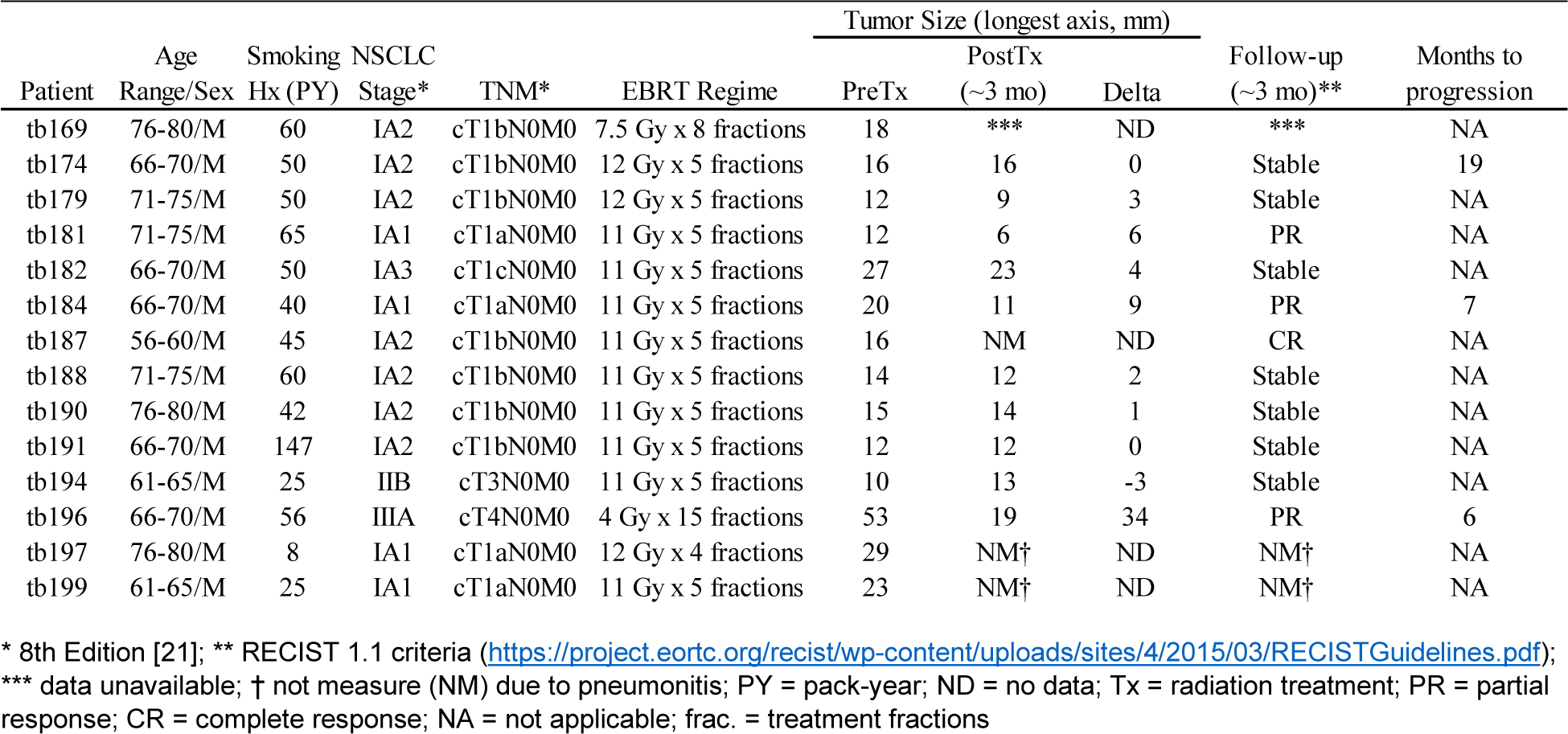
Patient demographics and clinical details.

### 2.2 Clinical measurements

We collected the following clinical data on NSCLC patients in this study which can be found in Table 1: smoking history (pack-years), NSCLC and TMN stage at diagnosis (8^th^ Edition [21]), treatment regime, tumor size (longest axis) at diagnosis and at ∼3 month follow-up imaging, RECIST tumor status at follow-up imaging (RECIST 1.1 criteria), and number of months from diagnosis to progression for up to 2-years of surveillance (until study completion).

### 2.3 Sample processing and DNA isolation

Blood draws (20-30 mL) were fractionated within 1 hour of collection for cfDNA isolation using a double spin method [22]. Briefly, blood plasma was isolated by first spinning whole blood at 1,000 x g for 10 min. at 4° C, separating the top plasma layer into ∼1-ml aliquots, spinning those aliquots at 15,000 x g for 10 min. at 4° C, and transferring the supernatant to cryovials for storage at −80° C. The buffy coat fraction was aliquoted (∼200 μl) after the initial spin and stored at −80°C. Cell-free DNA was isolated from 5-10 ml of plasma using Macherey-Nagel NucleoSnap (Macherey-Nagel GmbH & Co., Duren, Germany) and white blood cell (WBC)-derived DNA was extracted using QIAgen Blood and Tissue kits (QIAGEN, Redwood City, CA, USA). All DNA extractions were quantified using the Qubit 3 (Thermo Fisher Scientific, Waltham, MA, USA) and size distribution was confirmed using the Bioanalyzer 2100 (Agilent Technologies, Santa Clara, CA, USA). Prior to library preparation DNA isolated from WBCs was fragmented by sonication to ∼150 bp using a Covaris E220 (Covaris Inc., Woburn, MA, USA). Cell-free DNA was not sonicated prior to library preparation.

### 2.4 DIDA-Seq NGS library preparation, hybridization capture enrichment, and sequencing

Dual-index degenerate adaptor sequencing (DIDA-Seq) libraries were prepared as previously described [23] using a hybrid capture target enrichment panel adapted from the lung cancer-specific CAPP-Seq panel developed by Newman and colleagues (2014) [9]. Briefly, cell-free DNA or sonicated WBC DNA was end-repaired and A-tailed prior to a 15 min. ligation at 20° C to Illumina-compatible sequencing adaptor oligos containing p5 and p7 multiplexing and UMI sequences (IDT USA, Coralville, IA, USA) using KAPA HyperPrep Reagents (KAPA Biosystems, Capetown, South Africa). Ligated DNA was PCR amplified using Illumina library amplification primers for 8-10 cycles and enriched by 18-hour hybridization capture using the xGen Hybridization and Wash kit (IDT USA, Coralville, IA, USA) with biotinylated oligos targeting regions described by Newman and colleagues [9]. Post-capture sequencing libraries were further PCR amplified for 8-10 cycles. Next-generation sequencing was carried out on the Illumina NovaSeq S4 platform using paired-end, 150 bp reads.

### 2.5 Sequencing data processing and mutation calling workflow

Our mutation calling workflow was developed and optimized to identify single nucleotide variations (SNVs) and insertion-deletions (indels) as outlined in Fig. 2a and Supplementary Fig. S1 (see Supplementary Materials and Methods for a detailed description of this workflow).

**Fig. 2.**
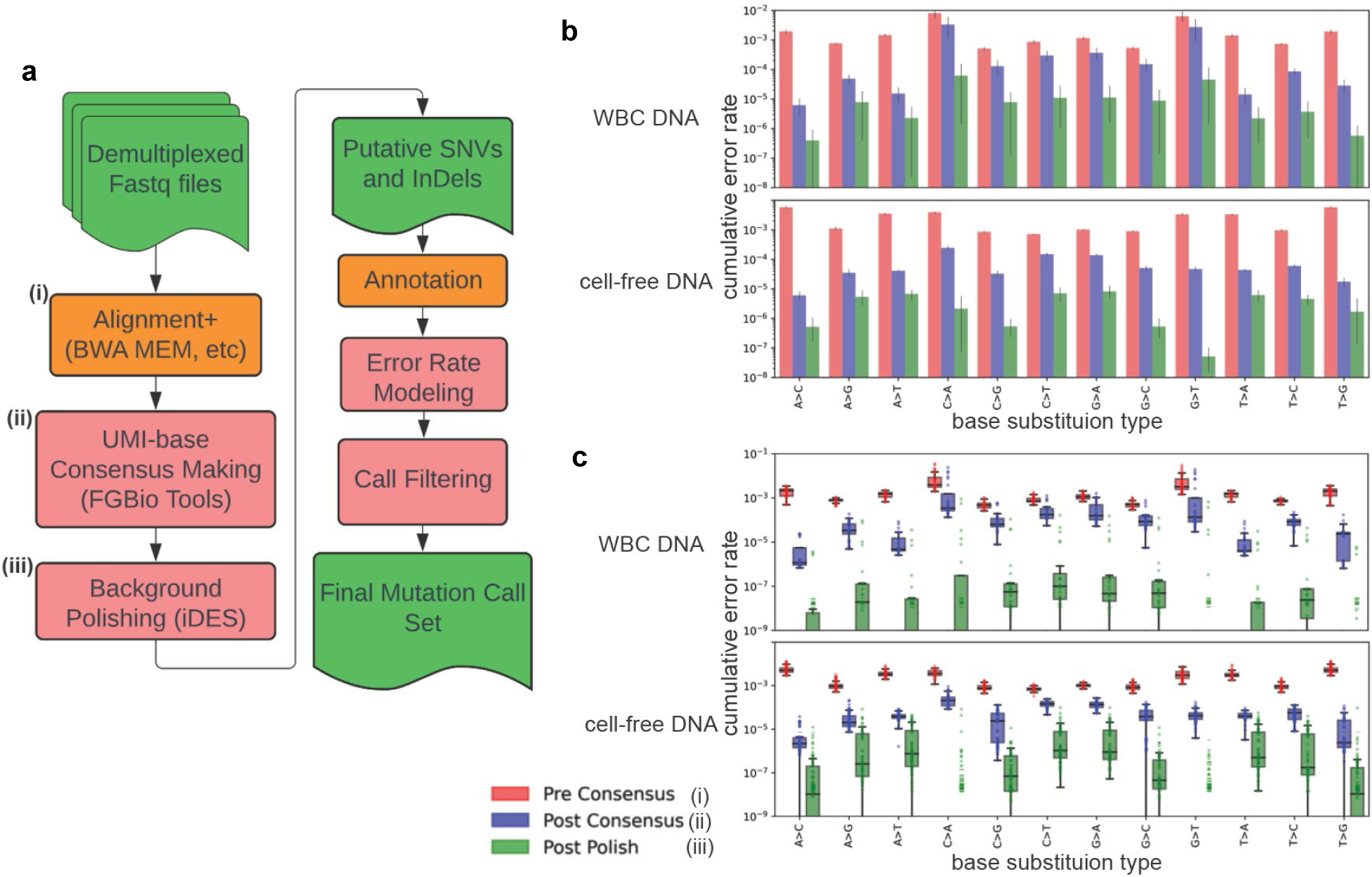
Condensed schematic of low-MAF *de novo* mutation calling pipeline (a) and associated error rate distributions (b and c) for cell-free DNA and WBC DNA at key stages (i-iii). A final mutation call set is generated for each input library (demultiplexed fastqs) after fastq alignment (**i**), consensus making (**ii**), background polishing (**iii**), and filtering steps (3^rd^ party tools indicated, see Methods and Supplementary Fig. S1 for details)). Red boxes indicate key points of pipeline optimization. Cumulative error rates (total alternate bases/total bases sequenced for each reference type, A, C, G, and T) are shown (**b**) with means and s.d. for each base substitution type for all cell-free DNA libraries and all WBC DNA libraries at three stages (**i-iii**) of error reduction pipeline shown in **a.** The distribution of cumulative error rates among libraries for each base substitution type is plotted (**c**) for three stages (**i-iii**) of the pipeline. In cases where there were no base substitutions of a given type in a library, a dash is plotted (in leu of a point) to indicate the total number of reference bases sequenced in a library [y = 1 / (total reference bases sequenced +1)]. This notation provides an estimate of the sampling limit when no errors were detected and is not intended to imply a lower limit of detection.

Briefly, raw fastq files were demultiplexed and written as unmapped bam files (ubam) using the *Demux* demultiplexing tool (Fulcrum Genomics, http://www.fulcrumgenomics.com) with no index mismatches allowed. Alignment and processing of ubam files was based on the GATK4 Best Practices Small-Variant Discovery Workflow (GATK, Broad Institute, Cambridge, MA, USA) with adaptations for consensus-making steps. These steps were carried out as depicted in Supplementary Fig. 1a using available tools and custom scripts [BWA-MEM v0.7.12 using hg38 [24], SAMtools v1.3 [25], and Picard tools v2.25.1 [26]]. The FGbio tool *CallMolecularConsensusReads* was used to identify PCR duplicates based on read-pair UMIs and mapping positions (UMI families) and *FilterConsensusReads* tool was used to filter consensus sequences. The steps and parameters implemented for these FGBio tools are described in Supplementary Materials and Methods. Consensus sequence bams were then filtered for tag-swapping events to eliminate false UMI families. To normalize for sequencing depth, consensus sequence bams were downsized prior to ctDNA detection analysis using *samtools subsample*. ctDNA dynamics were assessed with full-sized bam files (see Supplementary Materials and Methods for full details).

Background polishing of putative SNVs was done using the *Integrated Digital Error Suppression* (*iDES*) tools developed by Newman and colleagues [12] with default settings. Putative indels and SNVs mutation calls were annotated and subjected to filtering based on depth and coding status. We assumed that as the number of supporting reads for an alternate allele increases, the likelihood of error decreases. Therefore, we modelled the error rate for every base substitution type independently for each sequencing library as a function of the number of supporting mutant reads. Final calls were subjected to further filtering using these models and an error-rate threshold that was predetermined for ctDNA detection or measurement of ctDNA dynamics as described below and in Supplementary Materials and Methods.

### 2.6 Detection of gene fusions

We assessed full-sized and subsampled libraries for the presence of gene fusions within the target enrichment capture space using two software packages, *GeneFuse* and *FACTERA* [30, 31] using default settings.

### 2.7 Assessment of ctDNA detection rates

We assessed ctDNA detection rates before and after radiation exposure by comparing the number of patients with SNV or indel mutation calls passing filters primarily in BL and Tx1 blood draws. For each BL/Tx1 pair, the library with the greater mean depth was randomly subsampled to achieve parity depth as shown in Supplementary Fig. S2b. Each mutation call was assessed by the number of supporting alternate reads as described in Supplementary Materials and Methods. ctDNA was considered detectable if a putative mutation had sufficient supporting reads given a depth-specific error rate threshold previously determined to yield a false-positive rate (FPR) of zero (*i.e.,* no calls passing filters) in the healthy, cell-free DNA libraries (n=10) subsampled to match the depth of the library being analyzed (see Supplementary Materials and Methods and Supplementary Fig. S3). To assess the impact of multiple blood collections on detection rates, we considered all blood draws collected throughout treatment that had mean depths greater than BL/Tx1 parity depth for a given patient and then downsampled those libraries to achieve a comparable mean depth.

### 2.8 Characterization of mutations and ctDNA dynamics at BL and throughout treatment

For mutation characterization and ctDNA dynamics we considered all blood draws collected for each patient. Aligned bam files were not subsampled prior to mutation calling and filtering. For each library, we used the error rate threshold predicted by an exponential regression model of error rate as a function of mean depth to assess putative mutation calls based on the number of supporting reads (see Supplementary Materials and Methods and Supplementary Figs. S3 and S4). Mutation calls that were unique to draws other than baseline and Tx1 are indicated in Fig. 3 and Figs. 6a and 6b and were not included in the evaluation of ctDNA detection rates. All subsampled and full-sized bam files were subject to mutation calling and filtering using identical workflows as outlined above (see Supplementary Fig. S1).

**Fig. 3.**
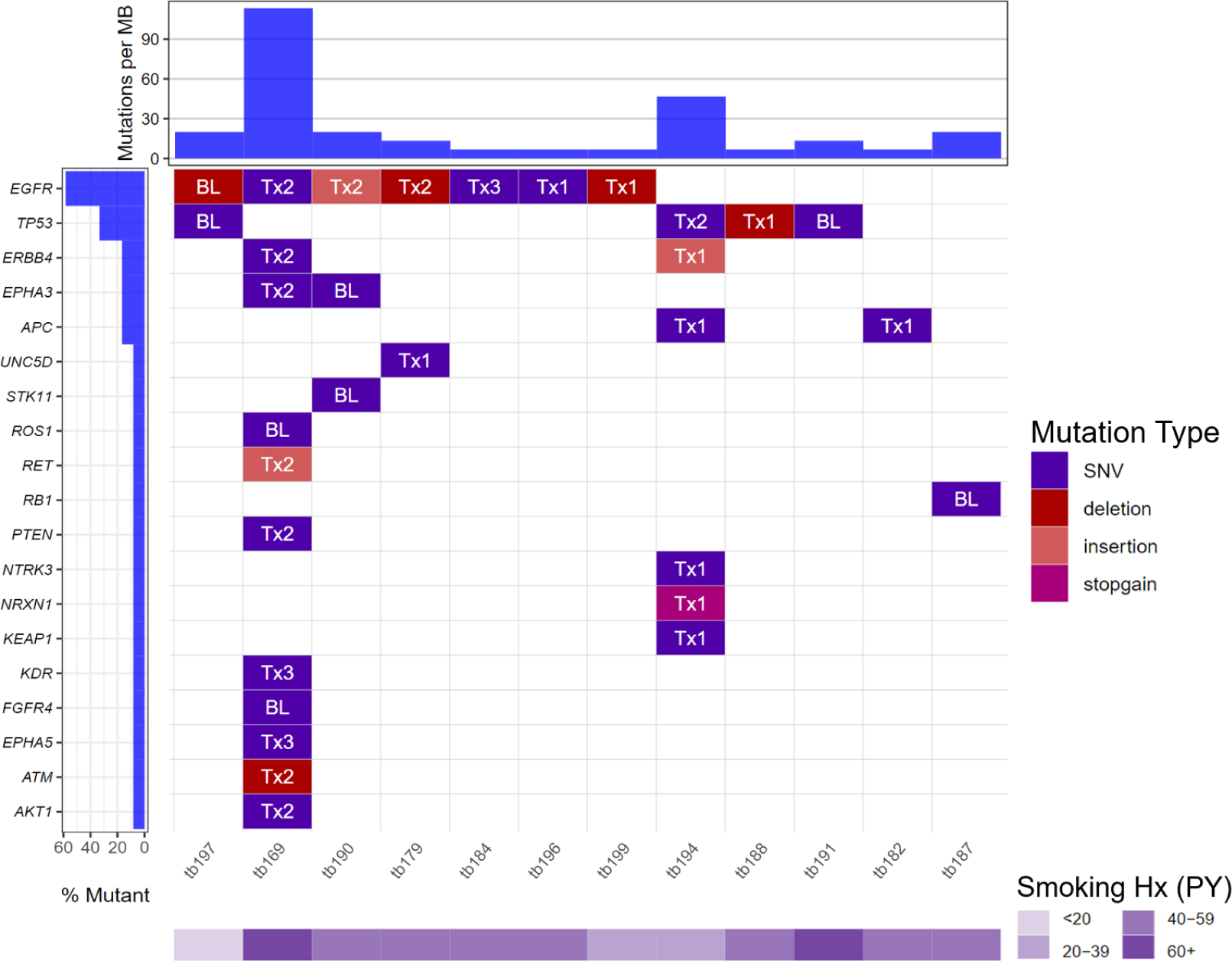
Non-synonymous mutations detected in cell-free DNA libraries collected at baseline and during radiation treatment of NSCLC patients. Waterfall plot showing single nucleotide variants (SNVs) and insertion/deletion events detected in cell-free DNA collected at baseline (BL) or on-treatment (Tx) as shown in Fig. 1b. The first draw in which the mutation was called is indicated. Mutation burden was calculated for the target enrichment capture space defined in the text. Smoking history is shown as pack-years (PY). Mutations appearing in this plot were called from full-sized bams and filtered as described in the text using probabilistic error rate thresholds calculated from library mean depth.

**Fig. 4.**
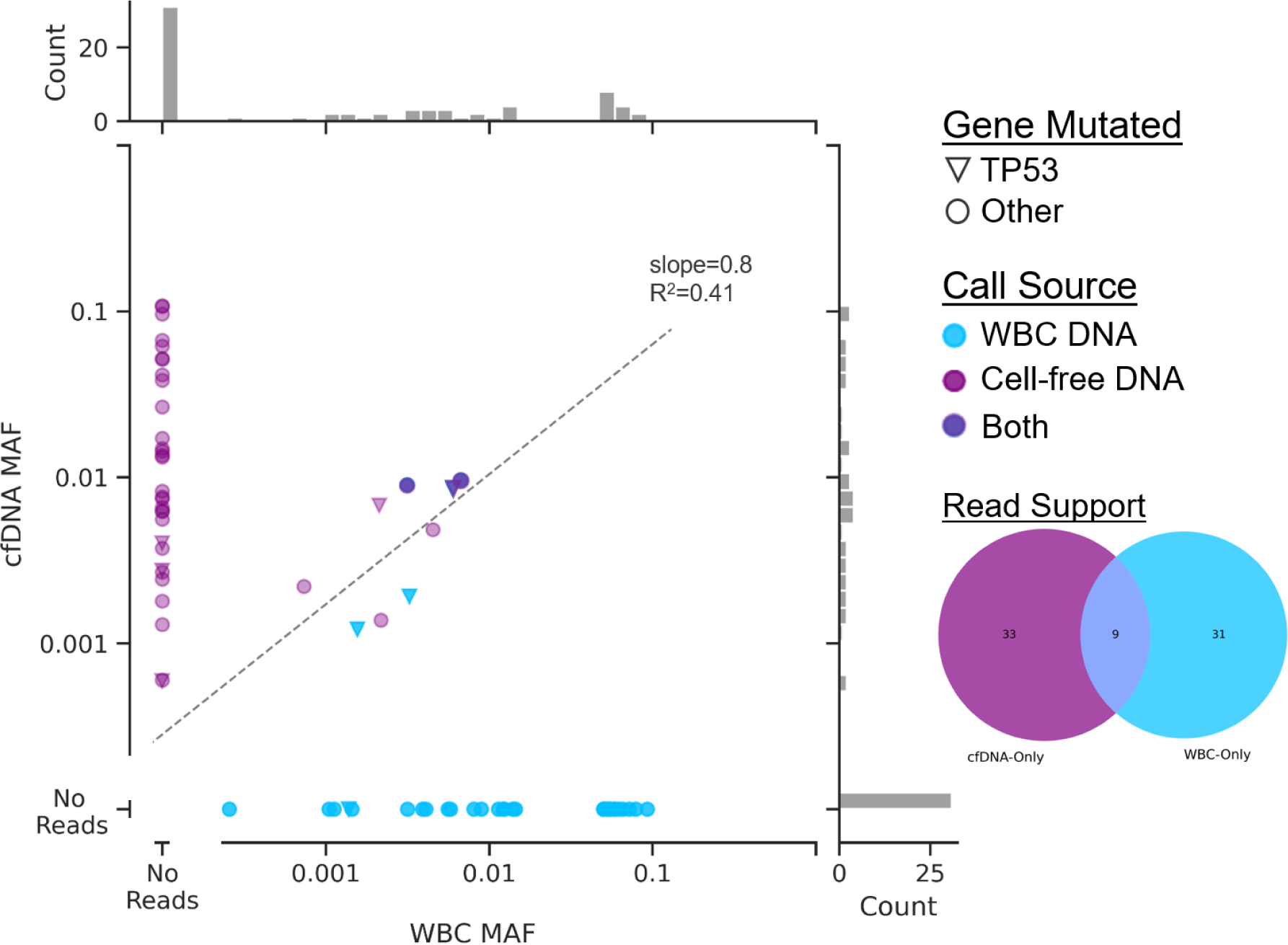
Prevalence of mutant reads in cell-free DNA and white blood cell (WBC)-derived DNA. Mutant allele fractions (MAF) are shown for mutation calls passing filters (without filtering for matched normal) in all cell-free DNA (lilac) and matched WBC DNA (blue) libraries. Mutations in TP53 are marked (▾). Mutations called in both tissue compartments are in dark blue (n=3, TP53, ERBB4, and NF1). Inset Venn diagram shows the distributions of read support in each tissue compartment for all mutations called (n=73 total, n=9 with reads in both tissue compartments).

**Fig. 5.**
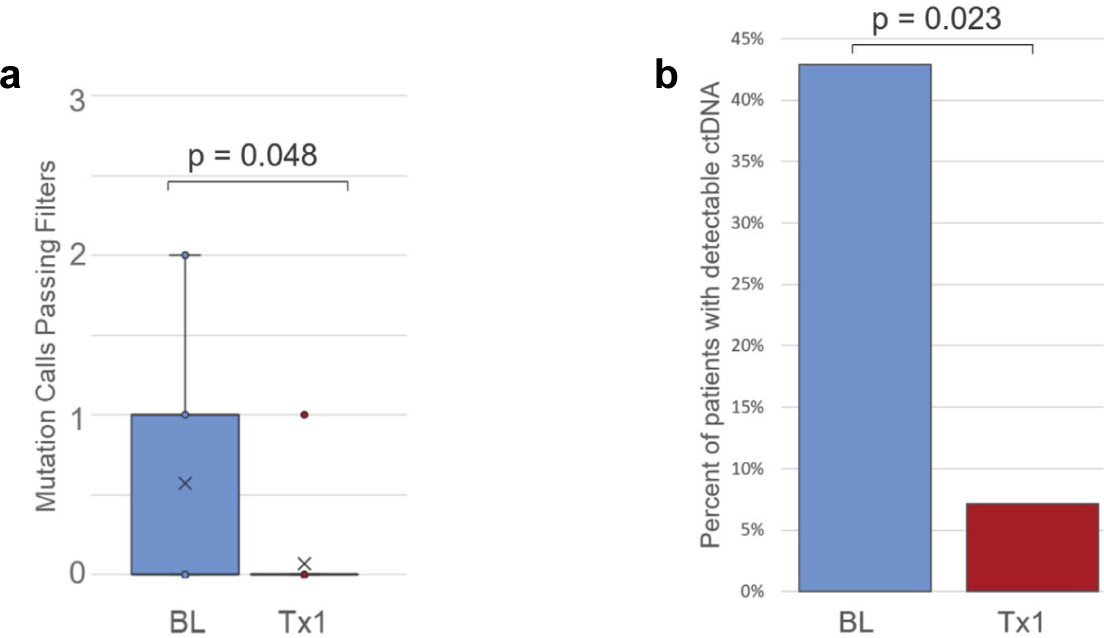
Comparison of final mutation call counts (a) and the proportion of NSCLC patients with detectable ctDNA (b) between baseline (BL) and first on-treatment (Tx1) blood draws (see Fig. 1a). (**a**) The number of ctDNA calls passing filters in BL and Tx1 draws, as reported in Table 2, was compared using a paired, two-tailed Student’s t-test (significance = 0.05). (**b**) The proportion of patients in Table 2 found to be ctDNA-positive between BL and Tx1 was compared using a two-tailed proportions z-score test (significance = 0.05).

**Fig. 6.**
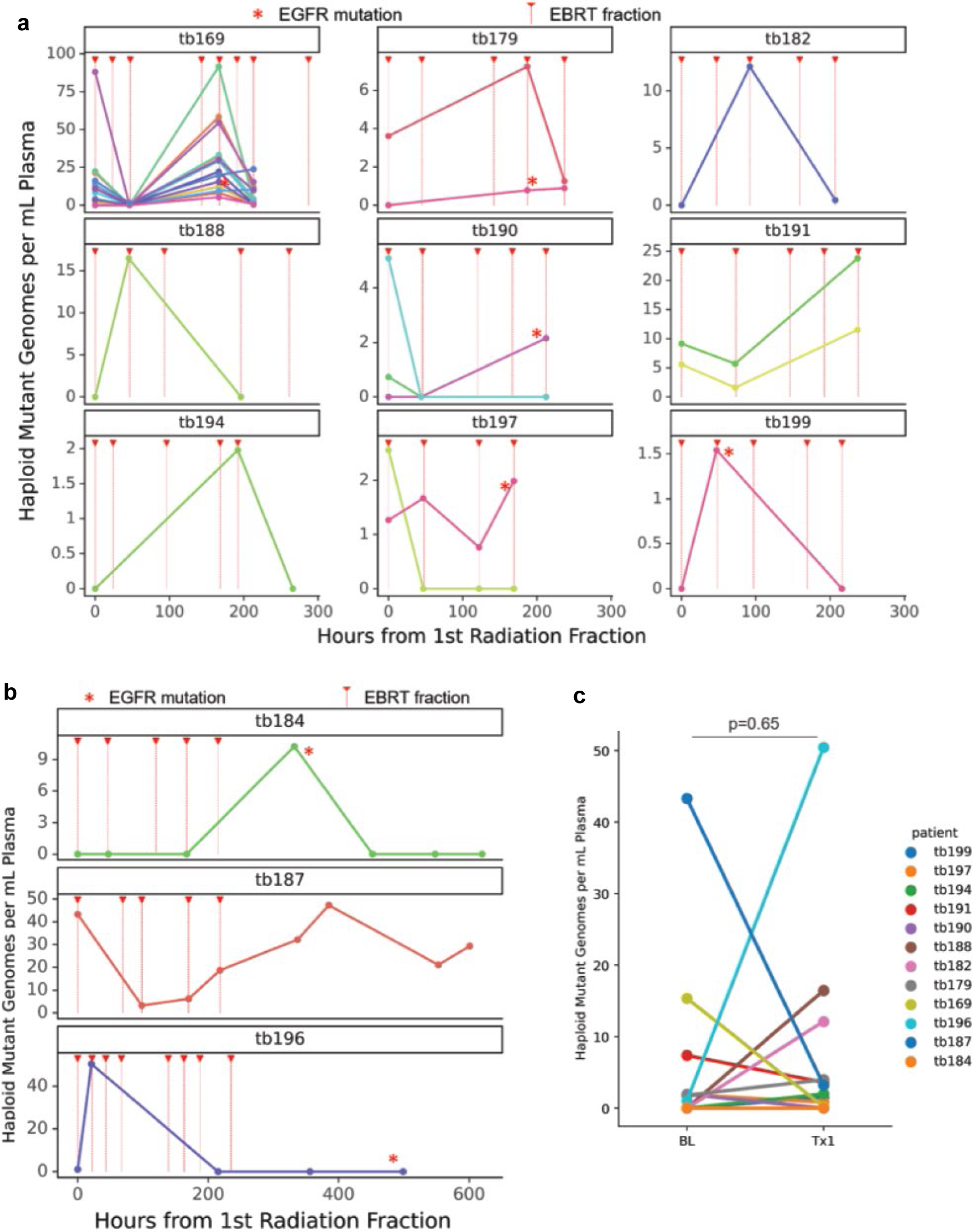
Dynamics of ctDNA abundance during fractionated external-beam radiation treatment (EBRT) and comparison of total ctDNA between baseline (BL) and the first on-treatment blood draw (Tx1). ctDNA abundance is shown for each patient during EBRT as mutant haploid genome equivalents per ml of plasma (hGE/ml plasma) for individual mutation calls passing filters at any time point in full-sized bam files. Red triangles and dashed lines indicate radiation treatment (see Table and Fig. 1b). Red asterisks indicate mutations in EGFR which were observed in patients tb169, tb184, and tb196. All blood draws that occurred on the same day as EBRT were taken ∼30-60 minutes prior to treatment. Nine patients had blood collected at BL (0 hours) and throughout treatment (**a**) and 3 patients also had draws collected after treatment completion (**b**). For each patient, read counts were summed for all mutation calls passing filters at BL and Tx1 and used to calculate total hGE/ml plasma (**c**) for statistical comparison between those time points (two-tailed paired t-test).

### 2.9 Sequencing error rate calculations

To measure the error reduction achieved by UMI-base consensus making and background polishing steps (Figs. 2b and 2c), we generated base call counts (pileups) at every position targeted by the capture enrichment panel with depth >1kX and MAF <20% for all libraries sequenced at three stages of our workflow: post-alignment, post-consensus making, and post-background polishing. We summed the base calls for every position and allele in each library and calculated the cumulative error rate for each base substitution type as the number of non-reference bases divided by the total number of reference and non-reference bases.

### 2.10 ctDNA abundance calculation

To account for variability in the amount of cell-free DNA in circulation at any given draw when measuring ctDNA abundance, we converted MAFs to haploid mutant genome equivalents per ml of plasma (hGE/ml plasma) as in our previous work [23]. MAFs were calculated using read counts acquired from full-sized bam files. Total ctDNA abundance at BL and Tx1 (Table 3) was calculated using summed alternate read counts and position depth for all mutations called at those time points.

**Table 2.** Number of mutation calls passing filters for each blood draw.^a^.

| Patient ID | NSCLC Stage | BL<sup>a</sup> | Tx1<sup>a</sup> | Tx2<sup>a</sup> | Tx3<sup>a</sup> | Tx4<sup>b</sup> | Tx5<sup>b</sup> | Tx6<sup>b</sup> | Tx7<sup>b</sup> |
| --- | --- | --- | --- | --- | --- | --- | --- | --- | --- |
| tb169 | IA2 | 2 | 0 | 12 | 3 | - | - | - | - |
| tb174 | IA2 | 0 | 0 | - | - | - | - | - | - |
| tb179 | IA2 | 0 | 0 | 1 | - | - | - | - | - |
| tb181 | IA1 | 0 | 0 | 0 | - | - | - | - | - |
| tb182 | IA3 | 1 | 0 | 0 | - | - | - | - | - |
| tb184 | IA1 | 0 | 0 | 0 | 1 | 0 | 0 | 0 | - |
| tb187 | IA2 | 1 | 0 | 0 | 0 | 1 | 1 | 0 | 0 |
| tb188 | IA2 | 0 | 0 | 0 | - | - | - | - | - |
| tb190 | IA2 | 1 | 0 | 1 | - | - | - | - | - |
| tb191 | IA2 | 2 | 0 | 0 | - | - | - | - | - |
| tb194 | IIB | 1 | 0 | 0 | - | - | - | - | - |
| tb196 | IIIA | 0 | 1 | 0 | 0 | 0 | - | - | - |
| tb197 | IA1 | 0 | 0 | 0 | 1 | - | - | - | - |
| tb199 | IA1 | 0 | 0 | 0 | - | - | - | - | - |
| **ctDNA Detection Rate** |  | **43%** | **7%** | **23%** | **60%** |  |  |  |  |
| **Cumulative Detection Rate** |  | **43%** | **50%** | **57%** | **71%** |  |  |  |  |

<sup>a</sup> BL or Tx1 libraries subsampled to achieve parity mean depth and using empirical error threshold such that the FPR of subsampled healthy controls was 0; <sup>b</sup> Libraries analyzed at full depth using probabilistic error threshold (resulting FPR of full-depth healthy controls of 10%); FPR = false positive rate; BL = baseline; Tx = on- or post-treatment.

**Table 3.** Aggregate mutant and reference read (Rd) counts for each patient at baseline (BL) and first on-treatment (Tx1) blood collections.

| Patient | Baseline Draw (BL)* |  |  | 1st On-Treatment Draw (Tx1)* |  |  | Delta |
| --- | --- | --- | --- | --- | --- | --- | --- |
|  | Mut Rd Count | Ref Rd Count | hGE/mL plas* | Mut Rd Count | Ref Rd Count | hGE/mL plas* |  |
| tb169 | 96 | 18696 | 15.4 | 2 | 71226 | 0.2 | -15.2 |
| tb179 | 6 | 15525 | 1.8 | 11 | 11376 | 4.0 | 2.2 |
| tb182 | 0 | 6061 | 0.0 | 8 | 1292 | 12.1 | 12.1 |
| tb184 | 0 | 5082 | 0.0 | 0 | 6235 | 0.0 | 0.0 |
| tb187 | 14 | 2269 | 43.3 | 1 | 1742 | 3.2 | -40.1 |
| tb188 | 0 | 1300 | 0.0 | 13 | 4778 | 16.5 | 16.5 |
| tb190 | 22 | 14774 | 1.9 | 0 | 10515 | 0.0 | -1.9 |
| tb191 | 36 | 10708 | 7.4 | 16 | 15610 | 3.6 | -3.7 |
| tb194<sup>†</sup> | 0 | 1074 | 0.0 | 6 | 12195 | 2.0 | 2.0 |
| tb196<sup>†</sup> | 1 | 2095 | 1.0 | 32 | 803 | 50.5 | 49.5 |
| tb197 | 6 | 18332 | 1.9 | 1 | 16470 | 0.8 | -1.1 |
| tb199 | 0 | 6439 | 0.0 | 2 | 8548 | 1.5 | 1.5 |

\* BL vs. Tx1 hGE/mL plasma, $p = 0.51$ (paired, two-tailed Student's t-test, sig. = 0.05); † stage II and III disease (all other patients were stage I, see Table 1); see Fig. 6c for associated plots.

### 2.11 Statistical methods, coding, and figure generation

For each putative mutation, the allele fractions and depths were compared between negative control and NSCLC libraries using the Overlap Method as described in Supplementary Materials and Methods and in previous work [23]. The resulting Weitzman Coefficient was used as the p-value in each comparison. We carried out all regression analyses using the *SciPy Stats* python package [32]. For ctDNA detection, the number of mutation calls passing filters was evaluated between BL and Tx1 draws using a paired, two-tailed Student’s t-test. The same method was used to compare ctDNA abundance between BL and Tx1 (Fig. 6C). We used a two-tailed proportions z-score test (significance = 0.05) to compare the rate of detection between BL and Tx1 draws.

For evaluation of clinical parameters and ctDNA measurements we carried out linear regression analysis on multiple predictors and outcomes using regression analysis to generate a Pearson correlation coefficient and found no significant linear correlation between each series. We were not powered to make statistical evaluations for categorical variables, such as tumor response (RECIST 1.1 Criteria) at 3-month follow-up, and progression status at study completion.

All custom scripts and executables were written in Python 3.10.6 (https://www.python.org/). All data figures were generated using Python *matplotlib* (v3.6.3, https://matplotlib.org/) and *seaborn* packages (v0.11.2, https://seaborn.pydata.org/), and R (v4.1.1, https://cran.r-project.org/) using the *GenVisR* package (v1.30.0, http://bioconductor.org/packages/release/bioc/html/GenVisR.html, [33]).

## 3. Results

### 3.1 Study design, cohort details and sample collection

Newly-diagnosed NSCLC patients were consented for this study (n=12, n=1, and n=1, stage I, II, and III, respectively) and samples were collected during fractionated EBRT (Fig. 1 and Table 1). Each blood draw coinciding with an EBRT fraction was acquired 30-60 minutes prior to receiving radiation on the day of treatment. We were unable to sample patients at matching intervals, consequently the first on-treatment blood sample (Tx1) was collected after a single fraction of EBRT in 8 patients, after two fractions in 4 patients, and after 3 or 4 fractions in the remaining 2 patients. All but 2 patients (tb174 and tb194) had at least one additional collection during treatment. Three patients also had blood collected at multi-day intervals for ∼2 weeks after receiving the final EBRT fraction (tb184, tb187, and tb196) to survey post-treatment ctDNA dynamics.

### 3.2 Development and optimization of low-MAF de novo mutation calling pipeline

Fractional abundance of ctDNA (i.e., MAF) in early-stage NSCLC is typically at or below the background error rate of traditional NGS and the sensitivity of conventional mutation calling pipelines [6, 7, 34, 35]. In order to detect ingle nucleotide variants (SNVs) and short insertion/deletion evens (indels) below 1% MAF without foreknowledge of tumor genotype, we combined our previously published DIDA-Seq UMI-based workflow [16, 23, 36] with the CAPP-Seq NSCLC selector panel and in-silico background error-suppression methods, previously published by Newman and colleagues [9, 12]. We developed a hybrid computational workflow and expanded our mutation call filtering approach to minimize false positives (Fig. 2a and Supplementary Fig. S1).

First, to more accurately construct consensus reads and better leverage base quality score data for downstream call filtering, we replaced our majority-rules consensus maker with several FGbio tools (Fulcrum Genomics, http://www.fulcrumgenomics.com), which generate and filter consensus reads in a base quality score-aware manner (Supplementary Fig. S2). To optimize our computational workflow and call filtering algorithm, we sequenced an average of 79.9 ng (range 40ng-70ng, s.d. 39.5ng) of cell-free DNA and ∼500ng of white blood cell (WBC) DNA isolated from 10-20 mL of blood collected from healthy adults (n=10) to a median on-target, single-stranded consensus sequence (SSCS) depth of 7535X (IQR: 6817X-8853X). To reduce the incidence of stereotypical errors introduced during library preparation, such as 8-oxogaunine conversion (G>T and C>A), we next employed the background polishing method for SNVs developed by Newman and colleagues [9]. In combination, these steps reduced the error rates of our sequencing data by 50- to 1000-fold (Fig. 2b). Notably, WBC libraries had slightly higher pre-consensus error rates for G>T/C>A substitutions, likely due to DNA sonication during library preparation, however, cell-free DNA libraries saw a greater reduction in these error rates from both mitigation methods.

The remaining non-synonymous SNVs or indels where annotated and filtered to suppress false positives using catalogued positions (exclusion lists). We omitted mutation calls if there were ≥2 supporting consensus sequences in the patient-matched WBCs to avoid calls resulting from clonal hematopoiesis. We assessed the position-specific background error of each call position using a Bayesian approach with the healthy control libraries as our prior. We found significant variability in the error rates across individual libraries (Fig. 2c) and assumed that this would confound a generalized approach for putative call assessment. Therefore, we developed an algorithm that modeled the likelihood of error as a function of the number of supporting reads. For each mutation the model predicts the minimum number of supporting SSCSs needed to exceed a predetermined background error rate given the base substitution type. Using the healthy control libraries, we determined these thresholds empirically as the error rates for which the number of mutations passing filter (*i.e.*, false-positive rate, FPR) was zero (see Supplementary Table ST2) or probabilistically from the on-target mean depth of the given library (see Supplementary Materials and Methods and Supplementary Fig. S4). We subjected all cell-free DNA and WBC DNA to this mutation calling workflow.

### 3.3 ctDNA-derived mutations detected in NSCLC patients and the impact of clonal hematopoiesis (CH)

The limit of detection of ctDNA using NGS is determined by the number of unique molecules sequenced at a given genomic position. Non-synonymous mutation calls from all NSCLC patient cell-free DNA libraries are shown in Fig. 3 and Supplementary Table S3. We detected 34 non-synonymous SNVs and indels in 19 genes across the enrichment capture space, with the majority in the NSCLC-associated EGFR and TP53 genes.

To assess the contribution of clonal hematopoiesis (CH)-derived variants to cell-free DNA, we compared the MAFs of WBC calls to the MAFs of ctDNA calls from all cell-free DNA libraries Fig. 4. We reasoned that CH-derived cell-free DNA would be present in plasma at similar fractions to WBC DNA, however, tumor-derived ctDNA would not be detectable in the WBC compartment. We found 97 mutation calls from cell-free DNA and WBC DNA (43 and 38, respectively) passing filters. Only 9 mutation calls had supporting reads in both compartments and 3 of those were called in both compartments. Mutations with supporting reads in both compartments had similar MAFs suggesting that they were derived from CH (slope=0.8, R^2^=0.41). The majority of calls did not have supporting reads in their corresponding tissue compartment. Shared mutations occurred in TP53 (n=4) and in genes not typically associated with CH (CSMD1, ERBB4, OR6F1, and NF1, n=4). This suggests that filtering ctDNA call sets by simply omitting mutations found in genes commonly associated with CH may not be sufficient to remove false positives arising from CH. These results highlight the value of sequencing patient-matched WBC DNA when calling low-MAF ctDNA mutations.

### 3.4 Detection rates of ctDNA decreased after EBRT

To test our hypothesis that sampling cell-free DNA during EBRT improves ctDNA detection rates, we considered the number of mutation calls passing filters at baseline (BL) and at the first on-treatment blood draw (Tx1). We accounted for the variability in depth between BL and Tx1 libraries in each patient by subsampling either bam file to a mean depth equal to the lower of the two (see Table 2; Supplementary Fig. S2). We subsampled the healthy control libraries to these same depths and determined the parameters such that FPR=0 in those libraries (Supplementary Table ST2). We found that the number of total calls passing filters across all patients decrease from BL in the Tx1 blood draw (p = 0.048; Fig. 5a). The proportion of patients with detectable ctDNA (*i.e*., ≥1 calls passing filters) also decreased significantly from 43% at BL to 7% at Tx1 (p=0.023).

When considering the cumulative ctDNA detection rate after multiple blood draws (*i.e.*, BL, Tx1, Tx2, and Tx3), despite decreased ctDNA detection rates overall, we found that the percentage of ctDNA positive patients increased from 43% (6 out of 14) at BL, to 71% (10 out of 14) by Tx3. These results underscore the value of multiple serial collections when assessing patients with very low ctDNA levels. Interestingly, we found that EGFR mutations were detectable in patients during EBRT but not at baseline (tb169, tb184, tb190, tb196, tb197, and tb199, see Fig. 3 and Table 2). Without multiple draws for each healthy control, it is unclear if these results are simply due to multiple sampling and unrelated to EBRT. We did not detect gene fusions in any patient at any time point using multiple software tools nor was the number of calls passing filters associated with mean library depth (see Supplementary Table ST5).

### 3.5 ctDNA dynamics during EBRT were mixed among NSCLC patients

To assess ctDNA dynamics in each patient during EBRT, we estimated the total number of mutant genomes per mL of plasma at every time point for all calls made at any time point using non-subsampled bam files. We set the error rate threshold for mutation call filtering to the value predicted by our probabilistic model using a given library’s mean depth. Notably, this approach yielded one false positive in our healthy controls (FPR = 10%) and a slightly lower BL detection rate (36%, 5/14), which increased to 93% after 3 draws were considered (see Supplemental Table ST4). We found that 5 patients had increased total ctDNA abundance at Tx1, while 6 patients had decreased levels. Eight patients had at least one post-BL draw with increased ctDNA levels (Fig. 6). Notably, 2 of the 3 patients with samples collected after completing EBRT had peak ctDNA abundance between 300-600 hours post initial radiation fraction (tb184 and tb187, Fig. 6b). When we considered only BL and Tx1 draws for all patients, we saw no significant difference in mean mutant hGE/ml plasma (p=0.65; Fig. 6c and Table 3). However, both patients with stage II and III disease were among those with increased ctDNA at Tx1 vs BL, which is consistent with previous work in more advanced NSCLC and metastatic breast cancer [16, 17].

### 3.6 ctDNA detection and abundance were not significantly associated with clinical observations

We analyzed multiple clinical parameters against ctDNA measurements at baseline and the first time point including: smoking history, NSCLC stage at diagnosis, longest tumor dimension at diagnosis and at 3-month follow-up, treatment response (RECIST 1.1 criteria at follow-up), and outcome at study completion (progression vs. non-progression at any point within ∼2 years). We found no significant differences among our comparisons.

## 4. Discussion

Disease stage at diagnosis is a critical factor in overall patient outcome and survival in NSCLC. However, as early-stage NSCLC is typically discovered, diagnosed, and treated from imaging alone, the incidence of overdiagnosis and treatment of non-cancerous nodules and other masses is likely underestimated [3, 37, 38]. Although liquid biopsy seems ideally suited as a companion diagnostic, low ctDNA detection rates and highly-variable ctDNA levels in early-stage NSCLC still pose a significant technical challenge.

There is evidence that radiation treatment of solid tumors can potentiate ctDNA abundance in human subjects and animal models, but not without significant variability among subjects, and it is typically more pronounced with more advanced disease [16, 17, 19, 20]. Yet it is unclear which factors, both biological and physiological, might drive this variability. In this study, we hypothesized that ctDNA detection and characterization might improve during radiation treatment. We therefore sought to characterize ctDNA before, during, and after radiation treatment in early-stage NSCLC. Contrary to previous findings, ctDNA detection rates in our cohort decreased between baseline and on-treatment draws. However, ctDNA abundance also varied between patients during treatment. Some of this variability was likely due to sampling error considering the low number of mutant reads detected in many of the libraries, but we cannot rule out potential biological sources either. Our probabilistic error thresholding approach yield 93% ctDNA detection rates after 4 draws in all NSCLC patients, however multiple draws from the same healthy control donor were not available. Further studies using multiple serial sampling of healthy individuals would help clarify this result.

This study was also hindered by asynchronous blood collection timing between patients, particularly for the first on-treatment draw. Asynchronous EBRT fractions might also impact ctDNA dynamics depending on the influence of cumulative radiation fractions on ctDNA shedding. Based on previous work, we believe earlier and more frequent sample collection, at 3-, 6-, or 12-hour intervals, would be more informative of the effects of EBRT on ctDNA abundance. In locally advanced NSCLC patients, Breadner and colleagues [17] showed increased ctDNA abundance over BL with peaks at 24- and 3-hours in patients receiving chemoRT or chemotherapy after ICI (no RT), respectively. ctDNA detection rates in these cohorts were 70% and 90% at baseline compared to 43% in our cohort. This is likely due to more advanced disease generating higher overall ctDNA abundance in the Breadner et al. cohorts. They also observed increased abundance in patients undergoing palliative fractionated RT alone at 36 and 60 hours after the initial fraction. Their cohorts received fractional doses ranging from 2 to 9 Gy per fraction whereas our cohort received fewer fractions at higher doses (mean = 11.3 Gy per fraction, range = 4-12 Gy), which may have impacted our measurements given our first post-baseline sampling was at a median of 47 hours. It may be that at higher fractional doses, cell death is more acute and ctDNA shedding more immediate. However, it is possible that chemotherapy is a more effective means of inducing ctDNA shedding than RT or that early-stage patients do not consistently shed ctDNA in the same way as locally advanced patients do; ctDNA detection in patients with smaller tumors depends on much more than assay sensitivity. It has also been observed that changes in ctDNA abundance during therapy may be predictive of treatment efficacy, where temporary increases in ctDNA during treatment followed by ctDNA clearance is associated with improved patient outcomes (see our review, [39]). Additional studies should be done to clarify these questions.

Our study was unable to validate our mutation calls with matched tumor biopsies due to lack of availability. Future studies in which solid tumor genotyping is done in conjunction with on-treatment ctDNA analysis could establish a ground truth of ctDNA shedding and dynamics during therapy. Moreover, any such studies in early-stage NSCLC patients should include larger, more gender diverse cohorts to improve statistical analysis. Doing more studies that utilize deeper sequencing, more patients, and earlier blood collection, might yet reveal some utility in sample collection during treatment as a companion diagnostic. As sequencing cost continues to plummet, multiple draws over the course of a patient’s care will become more feasible and routine, thereby improving the diagnostic confidence of liquid biopsy for guiding treatment.

## 6. Institutional Review Board and Informed Consent Ethics Statements

The study was conducted according to the guidelines of the Declaration of Helsinki and approved by the Institutional Review Board of Oregon Health & Science University (IRB# 10163, originally approved 19 October 2017).

Informed consent was obtained from all subjects involved in the study. Specifically, human specimens and data (including whole blood and clinical information) were prospectively acquired from healthy donors and study participants undergoing first-line external-beam radiation after diagnosis of NSCLC by imaging (n = 14, 12 x stage I, 1 x stage II, and 1 x stage III) after their informed written consent (Oregon Health & Science University Institutional Review Board Study #10163, first approved 19 October 2017). Subject numbers presented here and the identities of the associated participants were not known to anyone outside of our research group.

## 7. Data Availability Statement

All data produced in the present study are available upon reasonable request to the authors.

## Supplementary Materials and Methods

**Supplementary Fig. S1.**
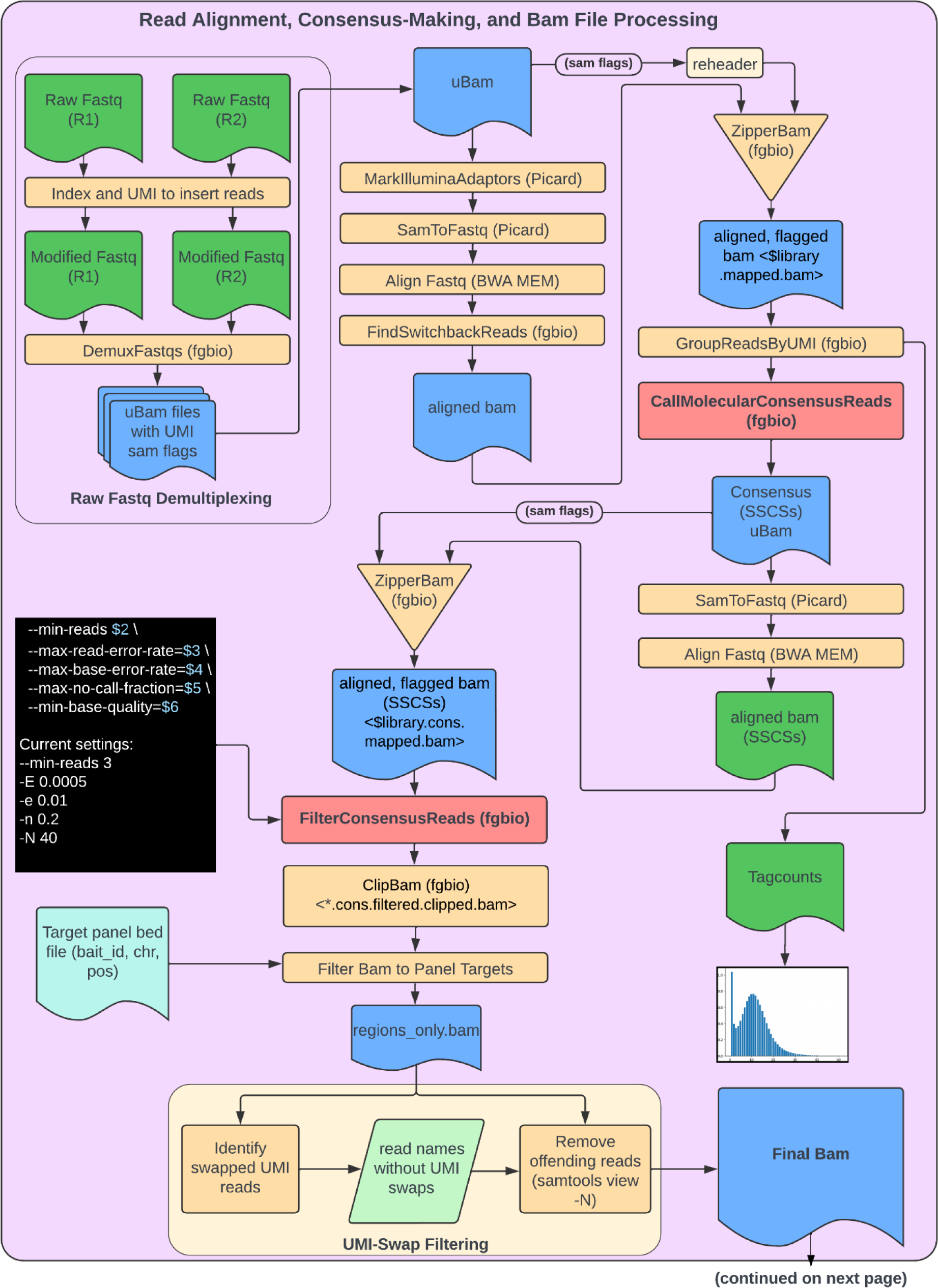
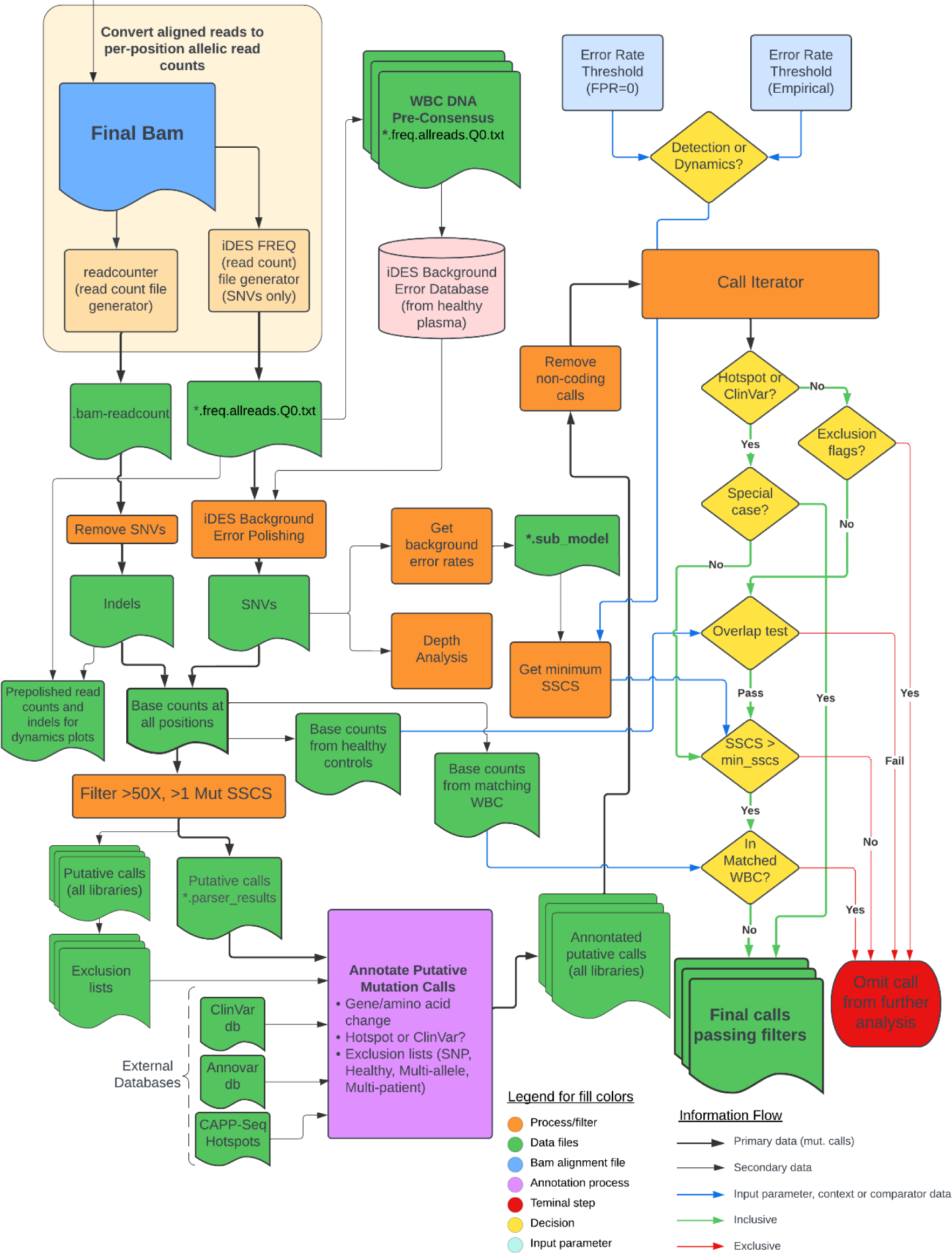
Complete schematic of low-MAF *de novo* mutation calling computational workflow. A diagram showing the data flow from sequencer output (raw paired-end fastqs) to final mutation calls (**a**). Files are indicated in blue and green; processes are indicated in tan and orange; major custom pipelines or algorithms are in grey, lilac and purple (i.e., fastq>bam, annotation script, and call filtering). Details of the annotation process are show in b. Third-party software is indicated when used and all other processes were done using custom scripts in Python 3.7.

**Supplementary Fig. S2.**
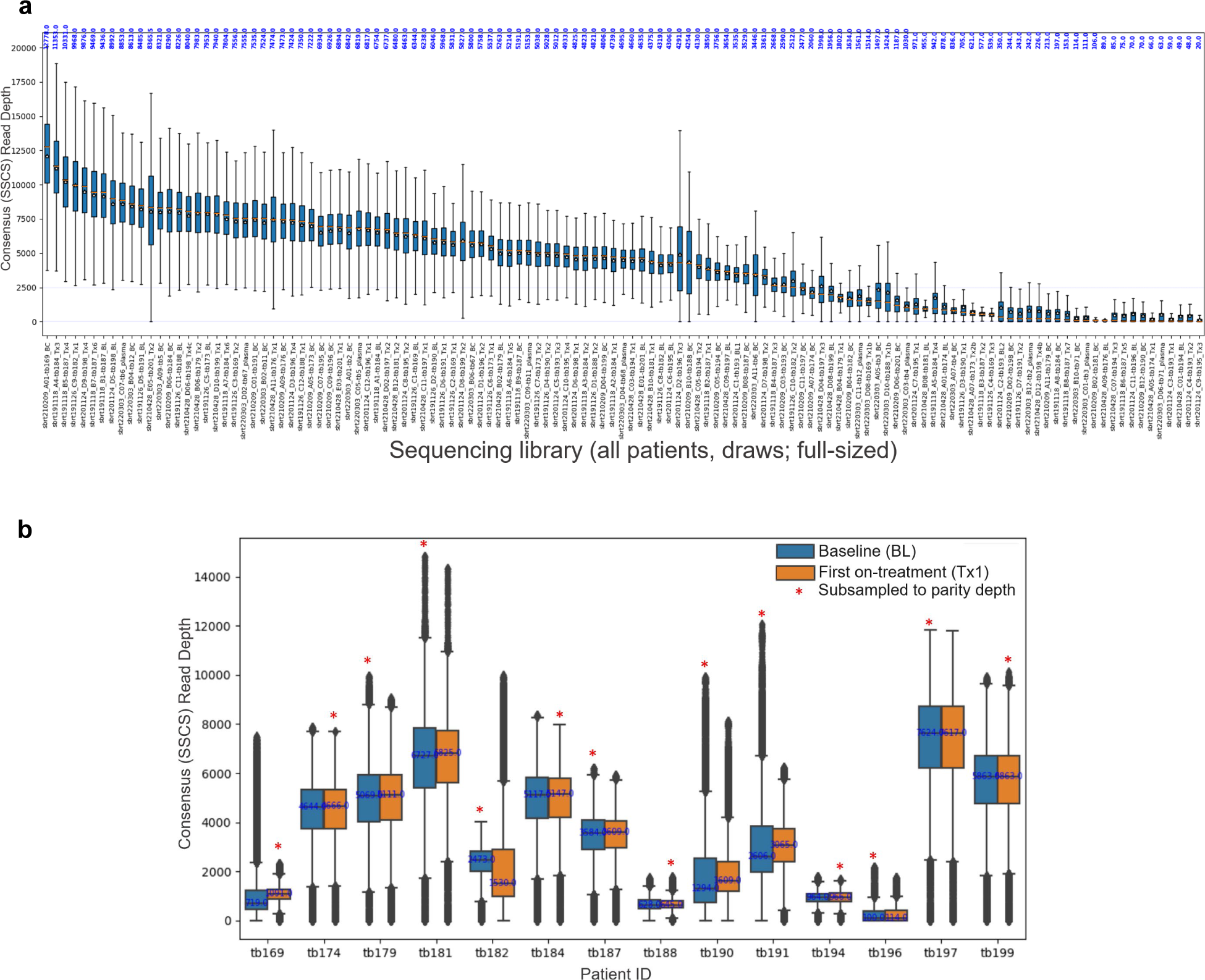
On-target depth distributions of sequencing libraries used in this study. Read counts were used to plot total coverage (depth) for all genomic positions covered in the CAPP-Seq [9] target enrichment hybridization capture panel (∼100kb). Read counts were derived from final full-sized bams (**a**) containing single-stranded consensus sequences (SSCSs) aligning to positions in the CAPP-Seq panel and represent unique input DNA molecules. Mean depths are shown as gold stars and medians are shown with orange dashes and values are annotated in blue. For each patient the depth distributions for the parity subsampled bams from baseline (BL) and first on-treatment (Tx1) blood draws are shown (**b**). Of the BL/Tx1 pairs, the bam which was subjected to subsampling to achieve parity mean depth is indicated with a red asterisk (see Supplementary Tables ST1 and ST2 for full-sized and subsampled mean depths for all libraries).

**Supplementary Fig. S3.**
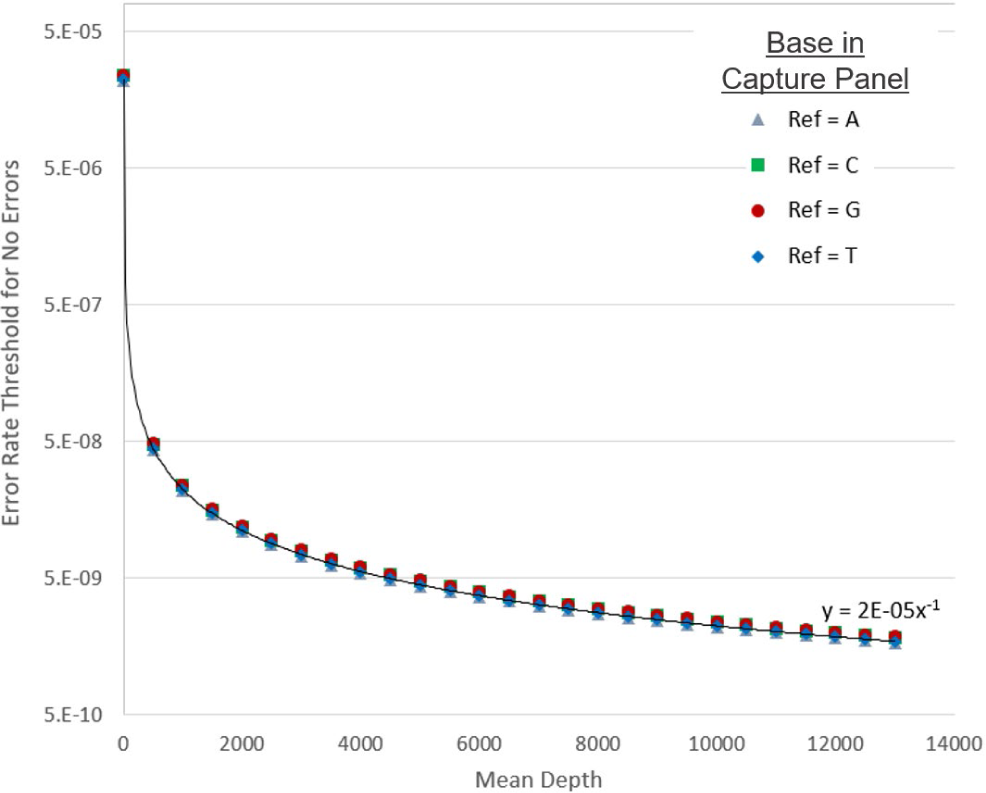
Relationship between the number of positions sequenced (defined as the maximum error rate tolerable, 1/total # of reference bases) and on-target mean depth. We used 1 over the total number of reference bases sequenced (y) to achieve a given mean depth (x) for each base type (A, C, G, and T) in the sequencing capture panel to calculate error rate thresholds used in ctDNA characterization and dynamics analysis. For example, at a library mean depth of 5000X, we expect ∼124M adenines to be sequenced in the capture panel, therefore, using the fit equation y=2E-5x^-1, where x=5000, we calculate that an error rate threshold of 1/124M or ∼8.05E-9 is needed to achieve less than 1 A>N errors in 124M adenines. This probabilistic approach was used to filter putative mutation calls in full-sized sequencing libraries (*i.e.*, not subsampled) and achieved a false positive rate of 10% when used to calculated error rate thresholds for filtering calls in healthy controls (n=10).

**Supplemental Fig. S4.**
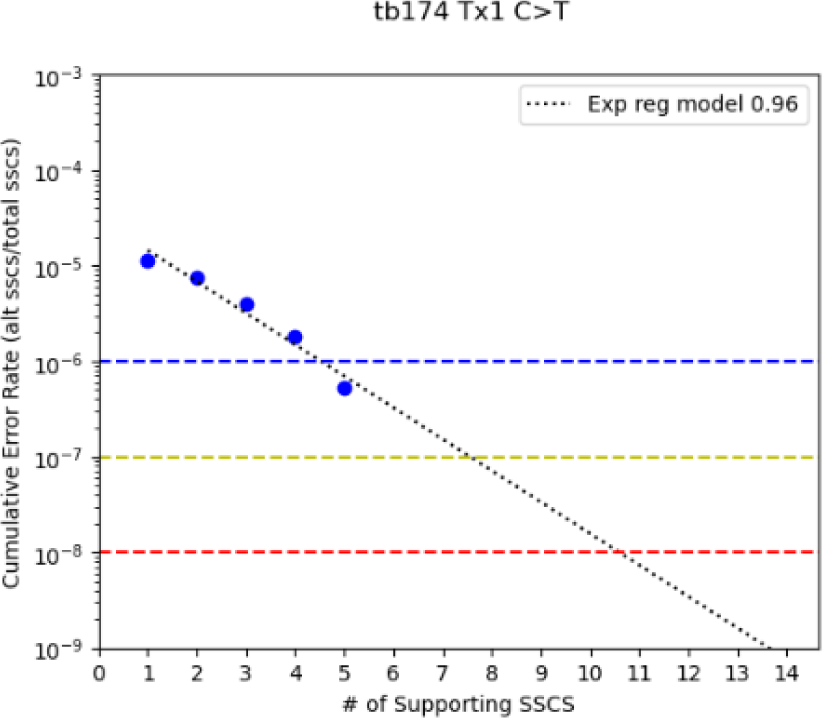
Example of the relationship between the error rate of a given base substitution type (*e.g.*, C>T, patient tb174 Tx1) and the number of supporting single-stranded consensus sequences (SSCSs) required for a given call to pass filters. For each library, the number of supporting reads (SSCSs) for a given base substitution type (A>C, A>G, …T>C, 24 total types) was counted across the entire library and plotted against the cumulative error rate, defined as the total alternate bases/total bases sequenced for each reference type, A, C, G, and T. We modeled this relationship by exponential regression and use this model to estimate the minimum number of supporting reads required to achieve a predefined error rate threshold, which was determined by either a probabilistic approach or empirically using depth-matched healthy controls. The intersection of the model [*i.e.*, x = [ln(y) – a]/b, where y=error rate and x=number of supporting reads] with the desired error rate threshold (solve for x) gives the minimum number of SSCSs needed to achieve that error rate. Colored dash lines reflect error rate thresholds of 1E-6 (blue), 1E-7 (yellow), and 1E-8 (red), which if specified, would require >5, >8, and >11 supporting SSCSs, respectively, for any C>T mutation called in this library.

**Supplementary Table ST1.**
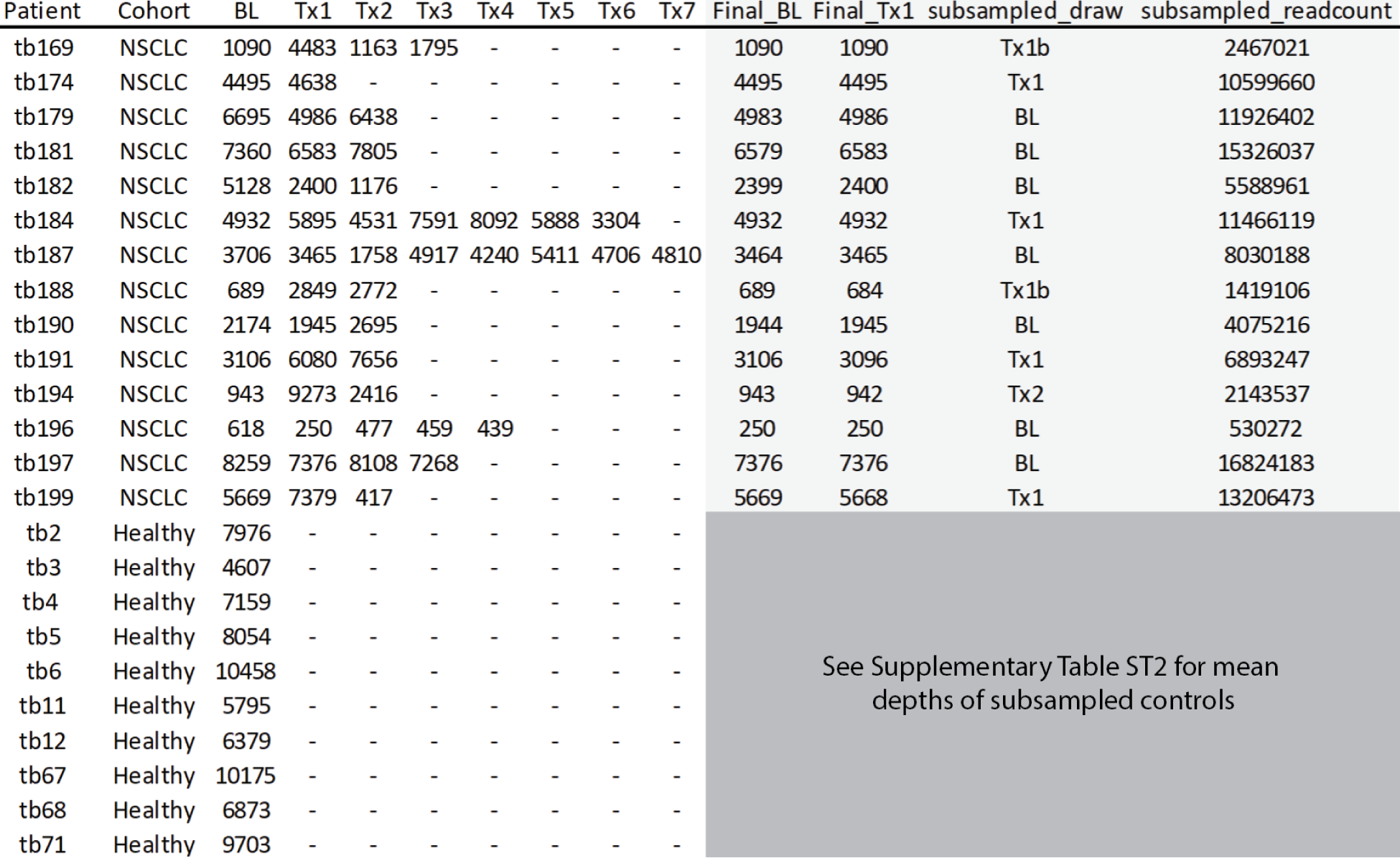
On-target mean consensus sequence depth of sequencing libraries. On-target mean consensus sequence depth of full-sized (not subsampled) bams and subsampled pairs (BL/Tx1), final parity depth, and read count (paired end).

**Supplementary Table ST2.** Library depth and calculated error rate thresholds and filtering parameters. Mean depth targets for healthy controls to achieve parity depth of patient’s BL/Tx1 pair. Empirically determined error rate thresholds are also shown for each patient target depth (parameters were provided to filtering algorithm in Supplementary Fig. S1).

|  |  | NSCLC Patient ID |  |  |  |  |  |  |  |  |  |  |  |  |  |
| --- | --- | --- | --- | --- | --- | --- | --- | --- | --- | --- | --- | --- | --- | --- | --- |
|  |  | tb169 | tb174 | tb179 | tb181 | tb182 | tb184 | tb187 | tb188 | tb190 | tb191 | tb194 | tb196 | tb197 | tb199 |
| Target DP<br>(mean DP of<br>NSCLC Patient): |  | 1090 | 4495 | 4983 | 6579 | 2399 | 4932 | 3464 | 684 | 1944 | 3096 | 942 | 250 | 7376 | 5668 |
| Healthy Patient ID | Healthy Patient Mean DP | Subsampled Depth |  |  |  |  |  |  |  |  |  |  |  |  |  |
| tb2 | 7976 | 1091 | 4479 | 5085 | 6600 | 2452 | 4916 | 3426 | 625 | 1911 | 3303 | 933 | 252 | 7410 | 5691 |
| tb3 | 4607 | 1078 | 4426 | 4607 | 4607 | 2421 | 4608 | 3384 | 618 | 1887 | 3262 | 921 | 248 | 4608 | 4608 |
| tb4 | 7159 | 1118 | 4591 | 5212 | 6775 | 2512 | 5039 | 3511 | 642 | 1957 | 3384 | 956 | 258 | 7159 | 5839 |
| tb5 | 8054 | 1103 | 4522 | 5133 | 6670 | 2474 | 4962 | 3458 | 632 | 1928 | 3333 | 943 | 255 | 7486 | 5750 |
| tb6 | 10458 | 1117 | 4581 | 5201 | 6757 | 2506 | 5029 | 3502 | 641 | 1953 | 3376 | 955 | 257 | 7584 | 5824 |
| tb11 | 5795 | 1075 | 4405 | 5000 | 5795 | 2411 | 4834 | 3367 | 617 | 1881 | 3245 | 919 | 248 | 5796 | 5601 |
| tb12 | 6379 | 1087 | 4459 | 5063 | 6397 | 2439 | 4894 | 3408 | 623 | 1900 | 3285 | 929 | 250 | 6379 | 5669 |
| tb67 | 10175 | 1083 | 4444 | 5046 | 6555 | 2432 | 4878 | 3399 | 621 | 1896 | 3276 | 925 | 250 | 7356 | 5650 |
| tb68 | 6873 | 1096 | 4485 | 5092 | 6614 | 2455 | 4923 | 3434 | 628 | 1915 | 3309 | 938 | 252 | 6874 | 5704 |
| tb71 | 9703 | 1104 | 4527 | 5138 | 6674 | 2478 | 4968 | 3460 | 633 | 1932 | 3335 | 944 | 254 | 7489 | 5752 |
| Average DS Depth |  | 1095.2 | 4491.9 | 5057.7 | 6344.4 | 2458.0 | 4905.1 | 3435.0 | 628.1 | 1916.1 | 3310.6 | 936.3 | 252.4 | 6814.1 | 5608.8 |
| Delta |  | 5.2 | -3.1 | 74.7 | -234.6 | 59.0 | -26.9 | -29.0 | -55.9 | -27.9 | 214.6 | -5.7 | 2.4 | -561.9 | -59.2 |
| Avg w/out mins |  | 1095.2 | 4491.9 | 5107.8 | 6663.6 | 2458.0 | 4938.1 | 3435.0 | 628.1 | 1916.1 | 3310.6 | 936.3 | 252.4 | 7465.1 | 5720.1 |
| Delta w/out mins |  | 5.2 | -3.1 | 124.8 | 84.6 | 59.0 | 6.1 | -29.0 | -55.9 | -27.9 | 214.6 | -5.7 | 2.4 | 89.1 | 52.1 |
| Prior target read count |  | 2589579 | 10636350 | 12075512 | 15112074 | 5816443 | 11674079 | 8129727 | 1483999 | 4532358 | 7835183 | 2157891 | 596237 | 17612077 | 13524254 |
| Average Reads per 1x |  | 2364 | 2368 | 2364 | 2268 | 2366 | 2364 | 2367 | 2363 | 2365 | 2367 | 2305 | 2362 | 2359 | 2364 |
| Empirical Error Threshold<br>Parameters where FPR=0 |  |  |  |  |  |  |  |  |  |  |  |  |  |  |  |
| --err_thr |  | 3E-08 | 3E-09 | 3E-10 | 3E-09 | 2E-09 | 5E-10 | 6E-09 | 3E-08 | 4E-08 | 5E-09 | 4E-08 | 5E-07 | 3E-09 | 1E-10 |
| --hs_err_thr |  | 1E-06 | 1E-07 | 1E-07 | 1E-07 | 1E-07 | 1E-07 | 1E-07 | 1E-06 | 1E-07 | 1E-07 | 1E-06 | 1E-06 | 1E-07 | 1E-07 |
| --tp53 |  | 1E-06 | 1E-07 | 1E-08 | 1E-07 | 1E-07 | 5E-08 | 1E-07 | 1E-06 | 1E-07 | 1E-07 | 1E-06 | 1E-06 | 1E-07 | 1E-07 |
| --exp_offset |  | 0 | 0 | 0 | 0 | 0 | 0 | 0 | 2 | 0 | 0 | 0 | 0 | 0 | 0 |

**Supplementary Table ST3.**
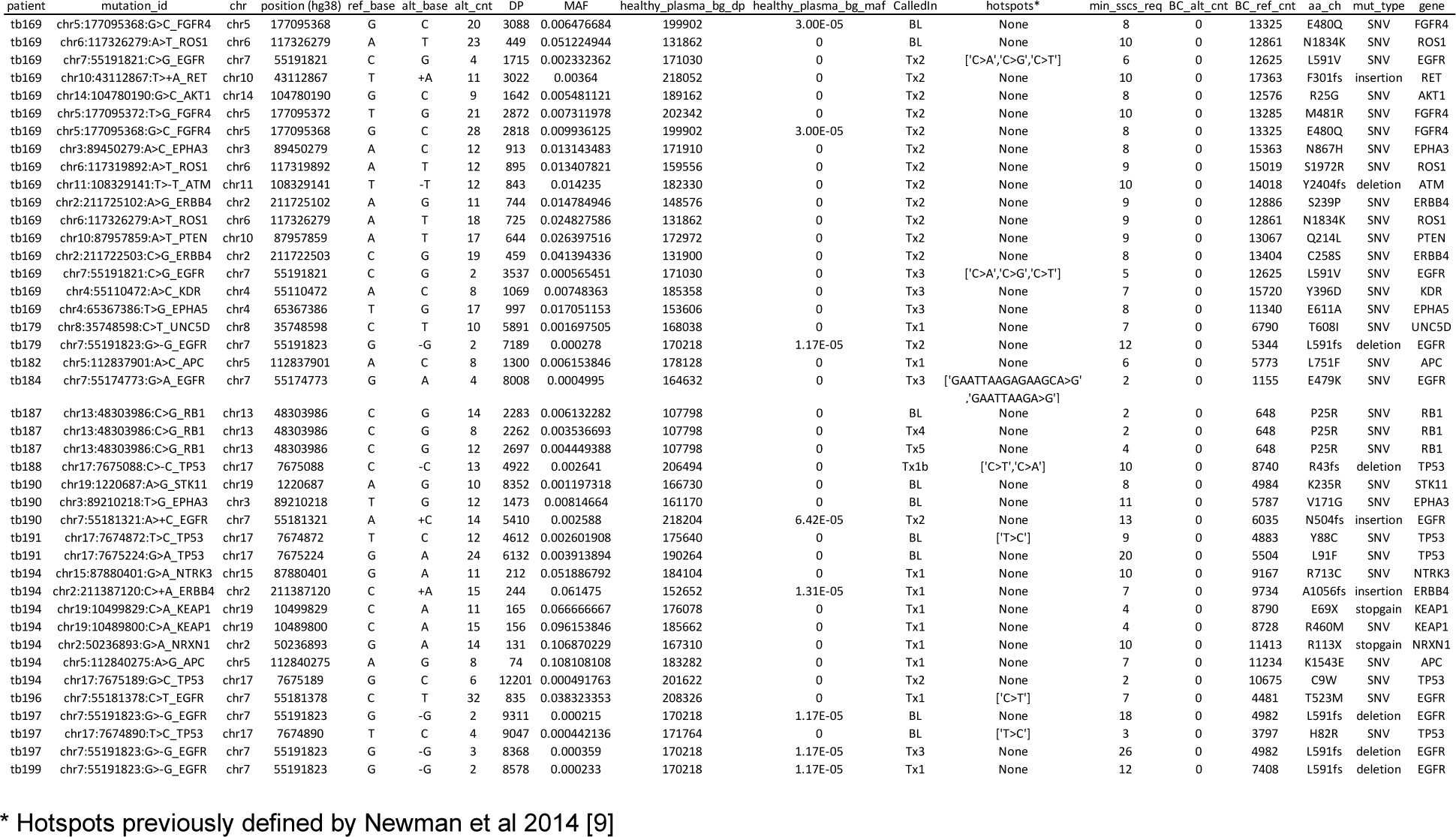
Final mutation calls passing filter from all time points.

**Supplementary Table ST4.** Number of mutation calls passing filters for each blood draw.

| patient | cohort | Blood Draw Library<sup>a</sup> |  |  |  |  |  |  |  |
| --- | --- | --- | --- | --- | --- | --- | --- | --- | --- |
|  |  | BL | Tx1 | Tx2 | Tx3 | Tx4 | Tx5 | Tx6 | Tx7 |
| tb169 | NSCLC | 2 | 0 | 12 | 3 | - | - | - | - |
| tb174 | NSCLC | 0 | 0 | - | - | - | - | - | - |
| tb179 | NSCLC | 0 | 1 | 1 | - | - | - | - | - |
| tb181 | NSCLC | 0 | 0 | 0 | - | - | - | - | - |
| tb182 | NSCLC | 0 | 1 | 0 | - | - | - | - | - |
| tb184 | NSCLC | 0 | 0 | 0 | 1 | 0 | 0 | 0 | - |
| tb187 | NSCLC | 1 | 0 | 0 | 0 | 1 | 1 | 0 | 0 |
| tb188 | NSCLC | 0 | 1 | 0 | - | - | - | - | - |
| tb190 | NSCLC | 2 | 0 | 1 | - | - | - | - | - |
| tb191 | NSCLC | 2 | 0 | 0 | - | - | - | - | - |
| tb194 | NSCLC | 0 | 6 | 1 | - | - | - | - | - |
| tb196 | NSCLC | 0 | 1 | 0 | 0 | 0 | - | - | - |
| tb197 | NSCLC | 2 | 0 | 0 | 1 | - | - | - | - |
| tb199 | NSCLC | 0 | 1 | 0 | - | - | - | - | - |
| (n) |  | 14 | 14 | 13 | 5 | 3 | 2 | 2 | 1 |
| ctDNA+ |  | 5 | 6 | 4 | 3 | 1 | 1 | 0 | 0 |
| Rate |  | 0.36 | 0.43 | 0.31 | 0.60 | 0.33 | 0.50 | 0.00 | 0.00 |
| Cumul. ctDNA+ |  | 5 | 11 | 11 | 13 | 13 | 13 | 13 | 13 |
| Cumul. Rate |  | 0.36 | 0.79 | 0.79 | 0.93 | 0.93 | 0.93 | 0.93 | 0.93 |
| tb2 | Healthy | 0 | - | - | - | - | - | - | - |
| tb3 | Healthy | 0 | - | - | - | - | - | - | - |
| tb4 | Healthy | 0 | - | - | - | - | - | - | - |
| tb5 | Healthy | 0 | - | - | - | - | - | - | - |
| tb6 | Healthy | 7 | - | - | - | - | - | - | - |
| tb11 | Healthy | 0 | - | - | - | - | - | - | - |
| tb12 | Healthy | 0 | - | - | - | - | - | - | - |
| tb67 | Healthy | 0 | - | - | - | - | - | - | - |
| tb68 | Healthy | 0 | - | - | - | - | - | - | - |
| tb71 | Healthy | 0 | - | - | - | - | - | - | - |
| (n) |  | 10 | - | - | - | - | - | - | - |
| ctDNA+ |  | 1 | - | - | - | - | - | - | - |
| Rate |  | 0.10 | - | - | - | - | - | - | - |

<sup>a</sup> All libraries were assessed at full-depth using error rate thresholds determined using our probabilistic approach and the library's mean coverage depth

**Supplementary Table ST5.**
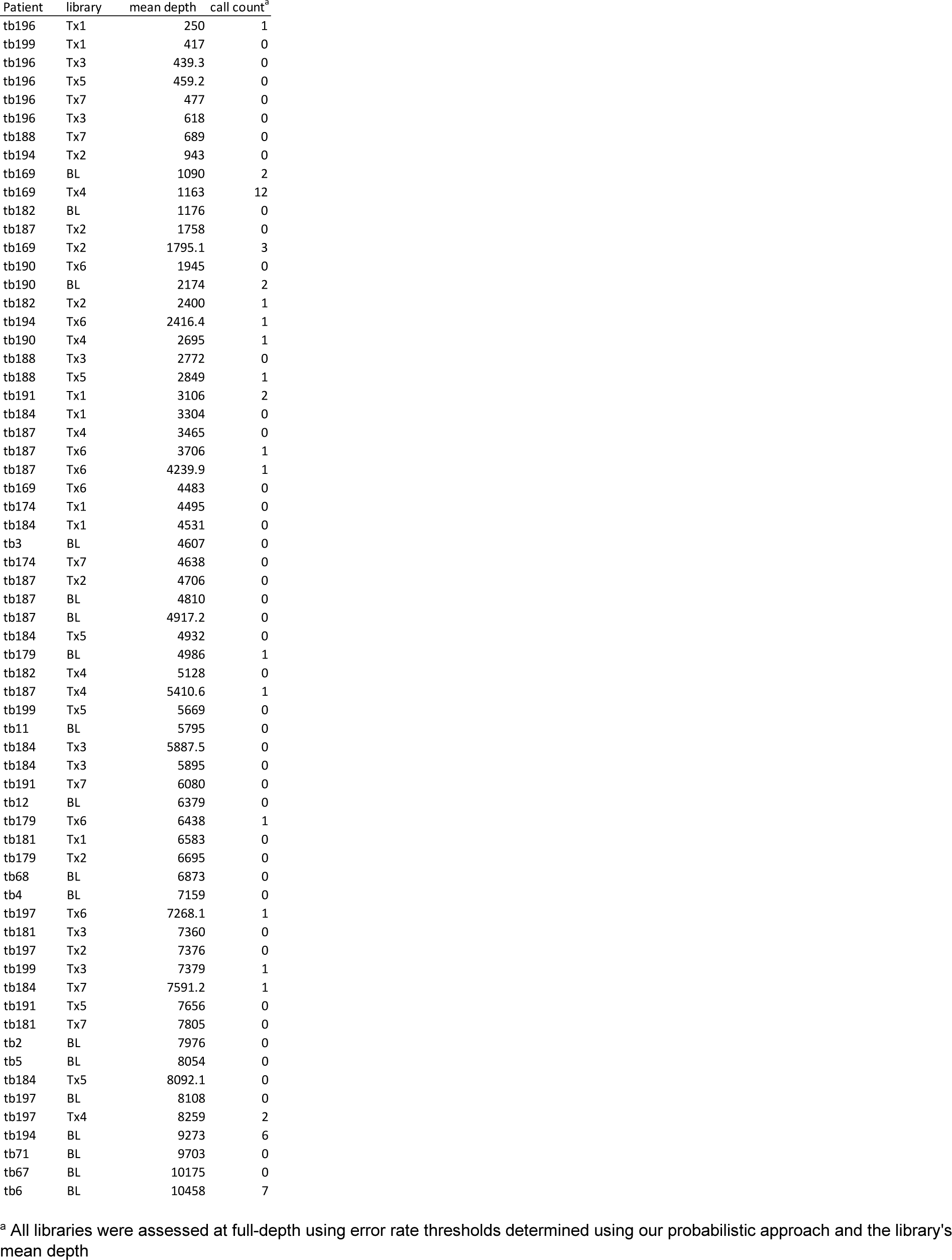
Library mean single-strand consensus sequence (SSCS) read depth and number of non-synonymous mutation calls passing filters.^a^.

### Description of FGBio tools used in consensus-making

The FGbio tools utilized in our consensus-making workflow, i.e., *CallMolecularConsensusReads* and FGbio *FilterConsensusReads*, are described by the developers here: http://fulcrumgenomics.github.io/FGbio/tools/latest/. Briefly, the *CallMolecularConsensusReads* tool first converts the base-quality scores (the MAPQ sam field) to error probabilities. Consensus reads are then called within UMI families using the individual base error probabilities to resolve inter-family base-call mismatches. Finally, a posterior error is assigned to each consensus base call as a consensus base quality score. The FGbio *FilterConsensusReads* tool was used to filter consensus sequences with the following settings: *--min-reads* 3, *--max-read-error-rate* 0.0005, *--max-read-error-rate* 0.01, *--max-no-call-fraction* 0.2, *--min-base-quality* 40.

### Identification and removal of UMI pseudo-families

Output consensus bam files were filtered for tag-swapping events (defined as two or more consensus sequences aligning to the same start and stop coordinates and possessing the same p5 <u>or</u> p7 UMI sequence <u>but not both</u>). A custom script was used to identify these events and output the consensus read with the lowest read error rate (the *cE* sam field) and the most PCR duplicates per pseudo-family (with random tie-breaks). The resulting consensus reads were written to a final bam file for error rate analysis, background polishing, and mutation calling. Base calls were extracted from bams at every genomic position of the target enrichment panel using the *bam-readcount* utility (https://github.com/genome/bam-readcount) unless otherwise specified (*i.e., iDES* background polishing).

### Background polishing of single-nucleotide variants (SNVs)

For SNVs, background polishing was carried out on read count files generated for each library using the *integrated digital-error suppression* (*iDES*) tools developed by Newman and colleagues [12] with default settings. We generated the *iDES* background error database used for subsequent background polishing with pre-consensus bams from the WBC DNA libraries, as they had the greatest impact on error reduction. Resulting read counts were later used to generate library-specific error rates for each substitution-type during call filtering. To generate a list of putative SNV calls, read count files were filtered to include only alternate base calls supported by at least 2 reads and 50X total coverage.

### Selection of putative insertions and deletions (indels)

Background error polishing was not carried out for detection of indels and we assumed that most recurrent artifacts causing indels (such as poorly mapped reads) would be present in multiple patient samples and therefore flagged during exclusion list generation and annotation steps later in our workflow. Instead, pre-polished read count files were filtered to exclude SNVs, and remaining indels were required to have ≥2 supporting reads and >50X coverage.

### Generation of exclusion lists and calculation of alternative allele background frequencies

We catalogued alternate alleles with the potential to give rise to false positives using the criteria described below (“hotspots” as defined by Newman *et al.* [9] were ignored). We combined read count files with putative SNVs and indels from plasma and WBC samples of NSCLC and healthy subjects, and then filtered them to exclude alternate alleles with <5 supporting reads. We catalogued likely SNPs (≥40% MAF) found in any library and all alternative alleles found in healthy cell-free DNA. Positions with alternate alleles present in multiple patients and with multiple alternative alleles found in a single patient were catalogued. All catalogued positions (*i.e.*, exclusion lists) were later used during annotation and filtering of putative mutation calls.

Alternate allele background frequencies were calculated for the “Overlap Method” test described below from all healthy cell-free DNA read count files. For each position in the target enrichment capture space, we recorded the number of supporting reads for all alleles (reference and alternate) and calculated the alternate allele fractions if present.

### Annotation of putative SNVs and indels

Read count files of putative SNVs and indels were annotated with amino-acid changes using the *Annovar* utility [27] (version 2019Oct24, https://annovar.openbioinformatics.org). Putative calls were also annotated with hotspot status and exclusion list flags catalogued above. We defined a hotspot as a mutation described in previous work by Newman and Colleagues [9] or reported as ‘pathogenic’ in the *ClinVar* [28] database (version 2023-01-21, https://ftp.ncbi.nlm.nih.gov/pub/clinvar/tab_delimited/variant_summary.txt.gz).

### Evaluation and filtering of putative SNVs and indels

We considered all non-synonymous putative SNVs and indels and removed all non-hotspot calls flagged as a SNP, present in healthy cfDNA, present in multiple patients, or with multiple alleles during annotation. To account for the variability in depth at each position in the target capture space, we used a Bayesian approach with the previously described Overlap Method [23, 29] when evaluating putative calls. Briefly, for each putative mutation, beta distributions were generated to test the null hypothesis that the putative MAF and the MAF of the same allele (if present) in healthy background frequency database arose from the same distribution. We omitted putative mutations if the calculated p-value exceeded 0.05 or 0.1, for non-hotspots and hotspots, respectively, in order to favor sensitivity over specificity for known or previous documented mutations. If the number of alternate read counts and total depth at the given position in the aggregate healthy controls was 0 and >50,000X, respectively, then the candidate mutation was retained. To mitigate contributions to cell-free DNA from clonal hematopoiesis, mutations having >1 supporting read in their matched WBC library were omitted from further consideration (this filter was not used for calls analyzed for clonal hematopoiesis, *Results 3.3*).

### Final mutation call evaluation by error probability as a function of number of supporting reads

To account for variability in the error rate of each base substitution type in each library, we evaluated putative mutations based on the number of supporting reads. We assumed that as the number of supporting reads for an alternate allele increases, the likelihood of error decreases. We modeled this behavior independently for each base substitution type in each library by exponential regression considering only positions having >1kX depth and with alternate alleles supported by <6 reads (see Supplementary Fig. S3). Putative SNVs having alternate read counts that were less than the minimum number required, as calculated from a given error rate threshold, were omitted from further consideration. For indels, we used the median value of all 3 substitution types matching the reference allele of the indel. Note that we evaluated hotspots using less stringent error rate thresholds (∼10-fold less) in order to give more statistical weight and flexibility in optimizing filtering.

### Calculation of the minimum error rate threshold for filtering mutation calls

We used error rate thresholds determined by either (a) fitting an exponential regression to the arrays *Y*, *X*, where *Y=1/[total reference base positions_A,C,T,G_ x mean depth]* and *X = mean depth = 1,2,3,…,13000*, and solving for y (*i.e.*, error rate threshold) given *X* (*i.e.*, mean depth of the call library) (Supplementary Fig. 4) or (b) empirically, where we calculated the error rate threshold for which the false-positive rate (FPR) was 0 (*i.e.,* no calls passing filters) in the healthy, cell-free DNA libraries subsampled to match the depth of the library being analyzed. The former approach was used for ctDNA characterization and dynamics (Figs. 3 and 6) and resulted in an FPR=0.1, and the latter approach was used for more stringent ctDNA detection rate analysis (Table 2 and Fig. 5) described below.

### Subsampling of bam files to normalize ctDNA detection rates at BL and Tx1

For comparison of ctDNA detection rates across time points from the same patient, final bam files were subsampled using the *samtools subsample* tool (Samtools v1.13) with a random seed and read output fraction. To assess the effect of EBRT on ctDNA detection rates, we limited our analysis to the BL and Tx1 draws. For each patient, we subsampled the bam file with the greater mean-depth, such that final mean-depth was within 10X of the lesser mean-depth library for that patient (see Supplementary Table ST1). Parity depth between bams was readily achieved for each patient by matching the total paired-end read count of the subsampled library to that of the target depth library. Healthy control libraries were each subsampled to match the parity mean depth of the patient being assessed. The resulting set of subsampled healthy controls was used to empirically determine an error rate threshold that achieved a false positive rate (FPR) of zero. Putative call filtering was then carried out using the respective threshold for each patient as appropriate.

## 8. References

[1] NIH, Surveillance, Epidemiology, and End Results (SEER) Program (www.seer.cancer.gov) SEER*Stat Database: Incidence - SEER Research Data, 8 Registries, Nov 2021 Sub (1975-2019) - Linked To County Attributes - Time Dependent (1990-2019) Income/Rurality, 1969-2020 Counties, National Cancer Institute, DCCPS, Surveillance Research Program, released April 2022, based on the November 2021 submission.

[2] H.J. de Koning, C.M. van der Aalst, P.A. de Jong, E.T. Scholten, K. Nackaerts, M.A. Heuvelmans, J.J. Lammers, C. Weenink, U. Yousaf-Khan, N. Horeweg, S. van’t Westeinde, M. Prokop, W.P. Mali, F.A.A. Mohamed Hoesein, P.M.A. van Ooijen, J. Aerts, M.A. den Bakker, E. Thunnissen, J. Verschakelen, R. Vliegenthart, J.E. Walter, K. Ten Haaf, H.J.M. Groen, M. Oudkerk, Reduced Lung-Cancer Mortality with Volume CT Screening in a Randomized Trial, N Engl J Med 382(6) (2020) 503–513.

[3] T. National Lung Screening Trial Research, T.R. Church, W.C. Black, D.R. Aberle, C.D. Berg, K.L. Clingan, F. Duan, R.M. Fagerstrom, I.F. Gareen, D.S. Gierada, G.C. Jones, I. Mahon, P.M. Marcus, J.D. Sicks, A. Jain, S. Baum, Results of initial low-dose computed tomographic screening for lung cancer, N Engl J Med 368(21) (2013) 1980–91.

[4] M. Jamal-Hanjani, G.A. Wilson, N. McGranahan, N.J. Birkbak, T.B.K. Watkins, S. Veeriah, S. Shafi, D.H. Johnson, R. Mitter, R. Rosenthal, M. Salm, S. Horswell, M. Escudero, N. Matthews, A. Rowan, T. Chambers, D.A. Moore, S. Turajlic, H. Xu, S.M. Lee, M.D. Forster, T. Ahmad, C.T. Hiley, C. Abbosh, M. Falzon, E. Borg, T. Marafioti, D. Lawrence, M. Hayward, S. Kolvekar, N. Panagiotopoulos, S.M. Janes, R. Thakrar, A. Ahmed, F. Blackhall, Y. Summers, R. Shah, L. Joseph, A.M. Quinn, P.A. Crosbie, B. Naidu, G. Middleton, G. Langman, S. Trotter, M. Nicolson, H. Remmen, K. Kerr, M. Chetty, L. Gomersall, D.A. Fennell, A. Nakas, S. Rathinam, G. Anand, S. Khan, P. Russell, V. Ezhil, B. Ismail, M. Irvin-Sellers, V. Prakash, J.F. Lester, M. Kornaszewska, R. Attanoos, H. Adams, H. Davies, S. Dentro, P. Taniere, B. O’Sullivan, H.L. Lowe, J.A. Hartley, N. Iles, H. Bell, Y. Ngai, J.A. Shaw, J. Herrero, Z. Szallasi, R.F. Schwarz, A. Stewart, S.A. Quezada, J. Le Quesne, P. Van Loo, C. Dive, A. Hackshaw, C. Swanton, T.R. Consortium, Tracking the Evolution of Non-Small-Cell Lung Cancer, N Engl J Med 376(22) (2017) 2109–2121.

[5] N. Cancer Genome Atlas Research, Comprehensive molecular profiling of lung adenocarcinoma, Nature 511(7511) (2014) 543–50.

[6] C. Abbosh, N.J. Birkbak, G.A. Wilson, M. Jamal-Hanjani, T. Constantin, R. Salari, J. Le Quesne, D.A. Moore, S. Veeriah, R. Rosenthal, T. Marafioti, E. Kirkizlar, T.B.K. Watkins, N. McGranahan, S. Ward, L. Martinson, J. Riley, F. Fraioli, M. Al Bakir, E. Gronroos, F. Zambrana, R. Endozo, W.L. Bi, F.M. Fennessy, N. Sponer, D. Johnson, J. Laycock, S. Shafi, J. Czyzewska-Khan, A. Rowan, T. Chambers, N. Matthews, S. Turajlic, C. Hiley, S.M. Lee, M.D. Forster, T. Ahmad, M. Falzon, E. Borg, D. Lawrence, M. Hayward, S. Kolvekar, N. Panagiotopoulos, S.M. Janes, R. Thakrar, A. Ahmed, F. Blackhall, Y. Summers, D. Hafez, A. Naik, A. Ganguly, S. Kareht, R. Shah, L. Joseph, A. Marie Quinn, P.A. Crosbie, B. Naidu, G. Middleton, G. Langman, S. Trotter, M. Nicolson, H. Remmen, K. Kerr, M. Chetty, L. Gomersall, D.A. Fennell, A. Nakas, S. Rathinam, G. Anand, S. Khan, P. Russell, V. Ezhil, B. Ismail, M. Irvin-Sellers, V. Prakash, J.F. Lester, M. Kornaszewska, R. Attanoos, H. Adams, H. Davies, D. Oukrif, A.U. Akarca, J.A. Hartley, H.L. Lowe, S. Lock, N. Iles, H. Bell, Y. Ngai, G. Elgar, Z. Szallasi, R.F. Schwarz, J. Herrero, A. Stewart, S.A. Quezada, K.S. Peggs, P. Van Loo, C. Dive, C.J. Lin, M. Rabinowitz, H. Aerts, A. Hackshaw, J.A. Shaw, B.G. Zimmermann, T.R. consortium, P. consortium, C. Swanton, Phylogenetic ctDNA analysis depicts early-stage lung cancer evolution, Nature 545(7655) (2017) 446–451.

[7] J.J. Chabon, E.G. Hamilton, D.M. Kurtz, M.S. Esfahani, E.J. Moding, H. Stehr, J. Schroers-Martin, B.Y. Nabet, B. Chen, A.A. Chaudhuri, C.L. Liu, A.B. Hui, M.C. Jin, T.D. Azad, D. Almanza, Y.J. Jeon, M.C. Nesselbush, L. Co Ting Keh, R.F. Bonilla, C.H. Yoo, R.B. Ko, E.L. Chen, D.J. Merriott, P.P. Massion, A.S. Mansfield, J. Jen, H.Z. Ren, S.H. Lin, C.L. Costantino, R. Burr, R. Tibshirani, S.S. Gambhir, G.J. Berry, K.C. Jensen, R.B. West, J.W. Neal, H.A. Wakelee, B.W. Loo, Jr., C.A. Kunder, A.N. Leung, N.S. Lui, M.F. Berry, J.B. Shrager, V.S. Nair, D.A. Haber, L.V. Sequist, A.A. Alizadeh, M. Diehn, Integrating genomic features for non-invasive early lung cancer detection, Nature 580(7802) (2020) 245–251.

[8] S. Waldeck, J. Mitschke, S. Wiesemann, M. Rassner, G. Andrieux, M. Deuter, J. Mutter, A.M. Luchtenborg, D. Kottmann, L. Titze, C. Zeisel, M. Jolic, U. Philipp, S. Lassmann, P. Bronsert, C. Greil, J. Rawluk, H. Becker, L. Isbell, A. Muller, S. Doostkam, B. Passlick, M. Borries, J. Duyster, J. Wehrle, F. Scherer, N. von Bubnoff, Early assessment of circulating tumor DNA after curative-intent resection predicts tumor recurrence in early-stage and locally advanced non-small-cell lung cancer, Mol Oncol 16(2) (2022) 527–537.

[9] A.M. Newman, S.V. Bratman, J. To, J.F. Wynne, N.C. Eclov, L.A. Modlin, C.L. Liu, J.W. Neal, H.A. Wakelee, R.E. Merritt, J.B. Shrager, B.W. Loo, Jr., A.A. Alizadeh, M. Diehn, An ultrasensitive method for quantitating circulating tumor DNA with broad patient coverage, Nat Med 20(5) (2014) 548–54.

[10] S. Avanzini, D.M. Kurtz, J.J. Chabon, E.J. Moding, S.S. Hori, S.S. Gambhir, A.A. Alizadeh, M. Diehn, J.G. Reiter, A mathematical model of ctDNA shedding predicts tumor detection size, Sci Adv 6(50) (2020).

[11] C. Abbosh, N.J. Birkbak, G.A. Wilson, M. Jamal-Hanjani, T. Constantin, R. Salari, J. Le Quesne, D.A. Moore, S. Veeriah, R. Rosenthal, T. Marafioti, E. Kirkizlar, T.B.K. Watkins, N. McGranahan, S. Ward, L. Martinson, J. Riley, F. Fraioli, M. Al Bakir, E. Gronroos, F. Zambrana, R. Endozo, W.L. Bi, F.M. Fennessy, N. Sponer, D. Johnson, J. Laycock, S. Shafi, J. Czyzewska-Khan, A. Rowan, T. Chambers, N. Matthews, S. Turajlic, C. Hiley, S.M. Lee, M.D. Forster, T. Ahmad, M. Falzon, E. Borg, D. Lawrence, M. Hayward, S. Kolvekar, N. Panagiotopoulos, S.M. Janes, R. Thakrar, A. Ahmed, F. Blackhall, Y. Summers, D. Hafez, A. Naik, A. Ganguly, S. Kareht, R. Shah, L. Joseph, A.M. Quinn, P.A. Crosbie, B. Naidu, G. Middleton, G. Langman, S. Trotter, M. Nicolson, H. Remmen, K. Kerr, M. Chetty, L. Gomersall, D.A. Fennell, A. Nakas, S. Rathinam, G. Anand, S. Khan, P. Russell, V. Ezhil, B. Ismail, M. Irvin-Sellers, V. Prakash, J.F. Lester, M. Kornaszewska, R. Attanoos, H. Adams, H. Davies, D. Oukrif, A.U. Akarca, J.A. Hartley, H.L. Lowe, S. Lock, N. Iles, H. Bell, Y. Ngai, G. Elgar, Z. Szallasi, R.F. Schwarz, J. Herrero, A. Stewart, S.A. Quezada, K.S. Peggs, P. Van Loo, C. Dive, C.J. Lin, M. Rabinowitz, H. Aerts, A. Hackshaw, J.A. Shaw, B.G. Zimmermann, C. Swanton, Corrigendum: Phylogenetic ctDNA analysis depicts early-stage lung cancer evolution, Nature 554(7691) (2018) 264.

[12] A.M. Newman, A.F. Lovejoy, D.M. Klass, D.M. Kurtz, J.J. Chabon, F. Scherer, H. Stehr, C.L. Liu, S.V. Bratman, C. Say, L. Zhou, J.N. Carter, R.B. West, G.W. Sledge, J.B. Shrager, B.W. Loo, Jr., J.W. Neal, H.A. Wakelee, M. Diehn, A.A. Alizadeh, Integrated digital error suppression for improved detection of circulating tumor DNA, Nat Biotechnol 34(5) (2016) 547–555.

[13] S.R. Kennedy, M.W. Schmitt, E.J. Fox, B.F. Kohrn, J.J. Salk, E.H. Ahn, M.J. Prindle, K.J. Kuong, J.C. Shen, R.A. Risques, L.A. Loeb, Detecting ultralow-frequency mutations by Duplex Sequencing, Nat Protoc 9(11) (2014) 2586–606.

[14] J. Moss, J. Magenheim, D. Neiman, H. Zemmour, N. Loyfer, A. Korach, Y. Samet, M. Maoz, H. Druid, P. Arner, K.Y. Fu, E. Kiss, K.L. Spalding, G. Landesberg, A. Zick, A. Grinshpun, A.M.J. Shapiro, M. Grompe, A.D. Wittenberg, B. Glaser, R. Shemer, T. Kaplan, Y. Dor, Comprehensive human cell-type methylation atlas reveals origins of circulating cell-free DNA in health and disease, Nat Commun 9(1) (2018) 5068.

[15] C. Boniface, K. Baker, C. Dieg, C. Halsey, R. Rahmani, M. Deffenbach, C. Thomas, G. Tormoen, N. Nabavizadeh, P. Spellman, Abstract PR05: Radiation-assisted Amplification Sequencing (RAMP-Seq): Evaluating the use of stereotactic body radiation therapy (SBRT) for enriching circulating tumor DNA in liquid biopsies, Clinical Cancer Research 26(11 Supplement) (2020) PR05–PR05.

[16] B.E. Johnson, A.L. Creason, J.M. Stommel, J.M. Keck, S. Parmar, C.B. Betts, A. Blucher, C. Boniface, E. Bucher, E. Burlingame, T. Camp, K. Chin, J. Eng, J. Estabrook, H.S. Feiler, M.B. Heskett, Z. Hu, A. Kolodzie, B.L. Kong, M. Labrie, J. Lee, P. Leyshock, S. Mitri, J. Patterson, J.L. Riesterer, S. Sivagnanam, J. Somers, D. Sudar, G. Thibault, B.R. Weeder, C. Zheng, X. Nan, R.F. Thompson, L.M. Heiser, P.T. Spellman, G. Thomas, E. Demir, Y.H. Chang, L.M. Coussens, A.R. Guimaraes, C. Corless, J. Goecks, R. Bergan, Z. Mitri, G.B. Mills, J.W. Gray, An omic and multidimensional spatial atlas from serial biopsies of an evolving metastatic breast cancer, Cell Rep Med 3(2) (2022) 100525.

[17] D.A. Breadner, M.D. Vincent, R. Correa, M. Black, A. Warner, M. Sanatani, V. Bhat, C. Morris, G. Jones, A. Allan, D.A. Palma, J. Raphael, Exploitation of treatment induced tumor lysis to enhance the sensitivity of ctDNA analysis: A first-in-human pilot study, Lung Cancer 165 (2022) 145–151.

[18] E.L. Chen, A.A. Chaudhuri, B.Y. Nabet, J.J. Chabon, D.J. Merriott, B.W. Loo, A.A. Alizadeh, M. Diehn, Analysis of Circulating Tumor DNA Kinetics during Stereotactic Ablative Radiation Therapy for Non-Small Cell Lung Cancer, International Journal of Radiation Oncology*Biology*Physics 102(3, Supplement) (2018) e676.

[19] G.M. Walls, L. McConnell, J. McAleese, P. Murray, T.B. Lynch, K. Savage, G.G. Hanna, D.G. de Castro, Early circulating tumour DNA kinetics measured by ultra-deep next-generation sequencing during radical radiotherapy for non-small cell lung cancer: a feasibility study, Radiat Oncol 15(1) (2020) 132.

[20] A. Rostami, M. Lambie, C.W. Yu, V. Stambolic, J.N. Waldron, S.V. Bratman, Senescence, Necrosis, and Apoptosis Govern Circulating Cell-free DNA Release Kinetics, Cell Rep 31(13) (2020) 107830.

[21] P. Goldstraw, K. Chansky, J. Crowley, R. Rami-Porta, H. Asamura, W.E. Eberhardt, A.G. Nicholson, P. Groome, A. Mitchell, V. Bolejack, S. International Association for the Study of Lung Cancer, A.B. Prognostic Factors Committee, I. Participating, S. International Association for the Study of Lung Cancer, B. Prognostic Factors Committee Advisory, I. Participating, The IASLC Lung Cancer Staging Project: Proposals for Revision of the TNM Stage Groupings in the Forthcoming (Eighth) Edition of the TNM Classification for Lung Cancer, J Thorac Oncol 11(1) (2016) 39–51.

[22] J.D. Merker, G.R. Oxnard, C. Compton, M. Diehn, P. Hurley, A.J. Lazar, N. Lindeman, C.M. Lockwood, A.J. Rai, R.L. Schilsky, A.M. Tsimberidou, P. Vasalos, B.L. Billman, T.K. Oliver, S.S. Bruinooge, D.F. Hayes, N.C. Turner, Circulating Tumor DNA Analysis in Patients With Cancer: American Society of Clinical Oncology and College of American Pathologists Joint Review, J Clin Oncol 36(16) (2018) 1631–1641.

[23] C. Boniface, C. Deig, C. Halsey, T. Kelley, M.B. Heskett, C.R. Thomas, Jr., P.T. Spellman, N. Nabavizadeh, The Feasibility of Patient-Specific Circulating Tumor DNA Monitoring throughout Multi-Modality Therapy for Locally Advanced Esophageal and Rectal Cancer: A Potential Biomarker for Early Detection of Subclinical Disease, Diagnostics (Basel) 11(1) (2021).

[24] H. Li, R. Durbin, Fast and accurate long-read alignment with Burrows-Wheeler transform, Bioinformatics 26(5) (2010) 589–95.

[25] P. Danecek, J.K. Bonfield, J. Liddle, J. Marshall, V. Ohan, M.O. Pollard, A. Whitwham, T. Keane, S.A. McCarthy, R.M. Davies, H. Li, Twelve years of SAMtools and BCFtools, Gigascience 10(2) (2021).

[26] Picard toolkit, Broad Institute, GitHub repository (https://broadinstitute.github.io/picard/) (2019).

[27] K. Wang, M. Li, H. Hakonarson, ANNOVAR: functional annotation of genetic variants from high-throughput sequencing data, Nucleic Acids Res 38(16) (2010) e164.

[28] M.J. Landrum, J.M. Lee, M. Benson, G.R. Brown, C. Chao, S. Chitipiralla, B. Gu, J. Hart, D. Hoffman, W. Jang, K. Karapetyan, K. Katz, C. Liu, Z. Maddipatla, A. Malheiro, K. McDaniel, M. Ovetsky, G. Riley, G. Zhou, J.B. Holmes, B.L. Kattman, D.R. Maglott, ClinVar: improving access to variant interpretations and supporting evidence, Nucleic Acids Res 46(D1) (2018) D1062–D1067.

[29] J.A. Montoya, G.F. P. D. González-Sánchez, Statistical inference for the Weitzman overlapping coefficient in a family of distributions, Applied Mathematical Modelling 71 (2019) 558–568.

[30] S. Chen, M. Liu, T. Huang, W. Liao, M. Xu, J. Gu, GeneFuse: detection and visualization of target gene fusions from DNA sequencing data, Int J Biol Sci 14(8) (2018) 843–848.

[31] A.M. Newman, S.V. Bratman, H. Stehr, L.J. Lee, C.L. Liu, M. Diehn, A.A. Alizadeh, FACTERA: a practical method for the discovery of genomic rearrangements at breakpoint resolution, Bioinformatics 30(23) (2014) 3390–3.

[32] P. Virtanen, R. Gommers, T.E. Oliphant, M. Haberland, T. Reddy, D. Cournapeau, E. Burovski, P. Peterson, W. Weckesser, J. Bright, S.J. van der Walt, M. Brett, J. Wilson, K.J. Millman, N. Mayorov, A.R.J. Nelson, E. Jones, R. Kern, E. Larson, C.J. Carey, I. Polat, Y. Feng, E.W. Moore, J. VanderPlas, D. Laxalde, J. Perktold, R. Cimrman, I. Henriksen, E.A. Quintero, C.R. Harris, A.M. Archibald, A.H. Ribeiro, F. Pedregosa, P. van Mulbregt, C. SciPy, SciPy 1.0: fundamental algorithms for scientific computing in Python, Nat Methods 17(3) (2020) 261–272.

[33] Z.L. Skidmore, A.H. Wagner, R. Lesurf, K.M. Campbell, J. Kunisaki, O.L. Griffith, M. Griffith, GenVisR: Genomic Visualizations in R, Bioinformatics 32(19) (2016) 3012–4.

[34] A.A. Chaudhuri, A.F. Lovejoy, J.J. Chabon, A. Newman, H. Stehr, D.J. Merriott, J.N. Carter, T.D. Azad, S. Padda, M.F. Gensheimer, H.A. Wakelee, J.W. Neal, B.W. Loo, A.A. Alizadeh, M. Diehn, Circulating Tumor DNA Analysis during Radiation Therapy for Localized Lung Cancer Predicts Treatment Outcome, International Journal of Radiation Oncology*Biology*Physics 99(2, Supplement) (2017) S1–S2.

[35] D.C. Koboldt, Best practices for variant calling in clinical sequencing, Genome Med 12(1) (2020) 91.

[36] T.M. Butler, C.T. Boniface, K. Johnson-Camacho, S. Tabatabaei, D. Melendez, T. Kelley, J. Gray, C.L. Corless, P.T. Spellman, Circulating tumor DNA dynamics using patient-customized assays are associated with outcome in neoadjuvantly treated breast cancer, Cold Spring Harb Mol Case Stud 5(2) (2019).

[37] B. Heleno, V. Siersma, J. Brodersen, Estimation of Overdiagnosis of Lung Cancer in Low-Dose Computed Tomography Screening: A Secondary Analysis of the Danish Lung Cancer Screening Trial, JAMA Intern Med 178(10) (2018) 1420–1422.

[38] J. Brodersen, T. Voss, F. Martiny, V. Siersma, A. Barratt, B. Heleno, Overdiagnosis of lung cancer with low-dose computed tomography screening: meta-analysis of the randomised clinical trials, Breathe (Sheff) 16(1) (2020) 200013.

[39] C.T. Boniface, P.T. Spellman, Blood, Toil, and Taxoteres: Biological Determinants of Treatment-Induced ctDNA Dynamics for Interpreting Tumor Response, Pathol Oncol Res 28 (2022) 1610103.

